# Single-cell transcriptome-wide association analysis reveals epithelial cell-specific genetic susceptibility to colorectal cancer

**DOI:** 10.64898/2026.10.29.26366877

**Authors:** Qing Li, Chao Li, Linshuoshuo Lyu, Jifeng Wang, Fangcheng Yuan, Eugene Jeong, Qingyuan Song, Chao Yan, You Chen, Jungyoon Choi, Shuai Xu, Yu Wang, Quanhu Sheng, Wanqing Wen, Martha J. Shrubsole, Qiuyin Cai, Xiao-Ou Shu, Guimin Gao, Bhuminder Singh, Zhijun Yin, Lori A. Coburn, Kristen K. Ciombor, Cathy Eng, Joshua A Bauer, Ulrike Peters, Keith T. Wilson, Wei Zheng, Ken S. Lau, Xingyi Guo

## Abstract

Bulk-tissue transcriptome-wide association studies (TWAS) have identified colorectal cancer (CRC) susceptibility genes, but cell-type-specific regulatory mechanisms remain poorly understood. Here, we performed single-cell TWAS integrating matched germline genotypes and single-cell transcriptomes from normal colorectal mucosa (381 donors) and polyps (84 donors), cell-type-specific chromatin accessibility, and GWAS from 132,261 cases and 1,752,398 controls. We identified 546 gene-cell-type associations implicating 347 candidate susceptibility genes across seven normal and nine polyp epithelial populations, including 213 previously unreported genes, with 74% exhibiting cell-type-specific associations. Integrated co-expression and ChIP-seq analyses revealed distinct TF-risk gene networks, highlighting GATA6, MAX and RAD21 in normal mucosa, and TFAP4- and CEBPB-associated networks in premalignant polyp populations. Further multi-omics analyses supported dysregulation of 233 genes (67%), while drug-target analyses identified 63 potentially druggable genes. DepMap and CRISPR–Cas9 analyses implicated 35 genes in CRC cell viability. These findings reveal cell-type-specific susceptibility mechanisms and prioritize candidate therapeutic targets for CRC prevention.

## Introduction

Genome-wide association studies (GWAS) have identified approximately 250 loci associated with colorectal cancer (CRC) susceptibility, providing important insights into the inherited genetic architecture of this malignancy^1–3^. However, translating these associations into the genes, cellular contexts and regulatory mechanisms underlying CRC susceptibility remains a major challenge. Most CRC-associated variants reside in noncoding genomic regions and are thought to influence disease risk by altering gene regulatory mechanisms, including transcription factor (TF) binding, chromatin accessibility, and cis-regulatory control of gene expression^4,5^. Transcriptome-wide association studies (TWAS), which integrate genetically regulated gene expression (GReX) with GWAS, offer a systematic approach to identify candidate susceptibility genes^6,7^. Previous CRC TWAS and other omics-based association studies^8–14^ have implicated more than 500 genes, advancing our understanding of the genetic basis of CRC susceptibility.

Despite these advances, previous studies have relied predominantly on GReX models trained in bulk normal colorectal tissue, potentially obscuring genetic regulatory effects restricted to specific epithelial populations or exhibiting opposing effects across cell types. Because regulatory variant activity is shaped by cell-type-specific chromatin accessibility and TF activity, bulk-derived models may overlook susceptibility genes and regulatory mechanisms operating within distinct epithelial states. Furthermore, genetic regulation in premalignant colorectal epithelialelium remains largely unexplored. CRC commonly develops from conventional adenomas or serrated polyps, which follow partially distinct molecular pathways toward malignancy^15–17^. The transition from normal mucosa to premalignant lesions involves extensive remodeling of epithelial differentiation programs, cellular composition, chromatin accessibility and transcriptional regulatory networks^15,18–20^. These changes may expose context-dependent genetic effects that are undetectable in normal tissue. Integrating both normal and premalignant epithelial populations into TWAS therefore strengthen the discovery of susceptibility genes and regulatory mechanisms associated with colorectal tumorigenesis.

Advances in population-scale single-cell RNA sequencing (scRNA-seq) coupled with germline genotyping now enable GReX modeling within defined cellular populations^11,21^. Incorporating cell-type-specific chromatin accessibility can further prioritize genetic predictors within biologically relevant regulatory regions, potentially improving model interpretability and the resolution of susceptibility-gene discovery^17^. Moreover, integrating cell-type-specific TWAS associations with TF-centered regulatory networks may reveal how inherited susceptibility genes are organized across normal and premalignant epithelial states^14,22,23^. Here, we developed a single-cell TWAS (scTWAS) framework integrating matched germline genotypes and single-cell transcriptomes from normal colorectal mucosa (381 donors) and polyps (84 donors), together with cell-type-specific chromatin accessibility data. We constructed GReX models across seven normal and nine polyp epithelial populations and integrated these models with European-ancestry and multi-ancestry CRC genetic association data comprising up to 132,261 cases and 1,752,398 controls. We further investigated cell-type-specific TF-susceptibility gene networks and integrated additional single-cell, spatial transcriptomic, bulk transcriptomic and proteomic evidence with drug-target annotations, cancer-dependency data and CRISPR-Cas9 perturbation analyses. This integrative framework resolves CRC susceptibility associations across distinct epithelial populations, identifies candidate TF-risk gene regulatory networks associated with premalignant states and prioritizes susceptibility genes with potential functional and translational relevance for CRC prevention.

## Results

### A cell-type-specific scTWAS framework

We developed a scTWAS framework integrating scRNA-seq, germline genotype and cell-type-resolved chromatin-accessibility data from normal colorectal mucosa and colorectal polyps (**Fig. 1a**; **Supplementary Table 1**). For each epithelial population, we aggregated single-cell expression profiles at the donor level and trained GReX models using variants located within cell-type-matched accessible chromatin regions^23^.

**Figure 1.**
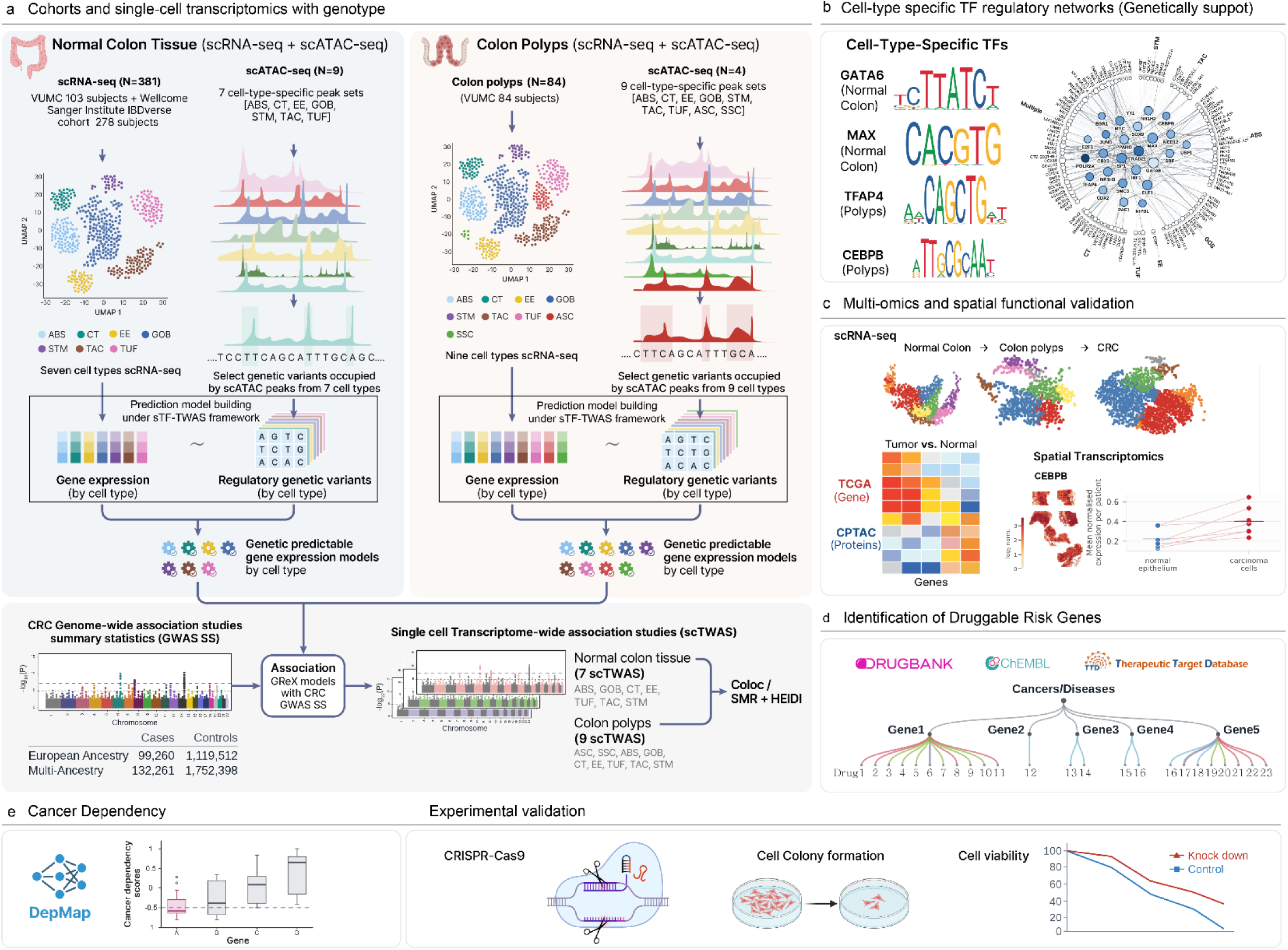
Study design and analytical framework. **a**, Cell-type-specific genetically regulated expression (GReX) modeling and single-cell transcriptome-wide association study (scTWAS). In normal colon and colorectal polyps, single-cell RNA-seq with matched germline genotype was integrated with cell-type-resolved scATAC-seq to define epithelial cell populations and identify genetic variants overlapping cell-type-specific accessible chromatin. These variants were used to train cell-type-specific GReX models from donor-level pseudobulk expression. The resulting models were integrated with colorectal cancer (CRC) GWAS summary statistics to perform scTWAS across seven normal-colon and nine polyp cell-type models, followed by colocalization and SMR–HEIDI analyses. b, Genetically supported cell-type-specific transcription-factor regulatory networks. CRC susceptibility-associated transcription factors were linked to scTWAS-identified risk genes to reconstruct cell-type-specific regulatory networks. c, Multi-omic and spatial functional validation. Risk genes and regulatory programs were evaluated across normal colon, colorectal polyps and CRC using single-cell and spatial transcriptomics, TCGA-COAD gene expression, CPTAC-COAD. d, Identification of druggable CRC risk genes. Therapeutic tractability was assessed by integrating DrugBank, ChEMBL, the Therapeutic Target Database and Open Targets. e, Cancer dependency and experimental validation. Candidate genes were evaluated using DepMap cancer-dependency data and experimentally tested by CRISPR–Cas9 perturbation followed by colony-formation and cell-viability assays.

For normal colorectal mucosa, we analyzed matched scRNA-seq and genotype data from 381 individuals, comprising two cohorts of 103 participants from the Human Tumor Atlas Network (HTAN) COLON MAP^24^ and Gut Cell Atlas studies ^25^ and 278 from the Wellcome Sanger Institute IBDverse cohort^21^ (**Fig. 1a; Supplementary Table 2**). We built GReX models for each of seven epithelial populations: absorptive (ABS), crypt-top (CT), enteroendocrine (EE), goblet (GOB), stem (STM), transit-amplifying (TAC) and tuft (TUF) cells (**Extended Data Fig. 1a, 1b**). Annotations from the COLON MAP normal-colon dataset served as the reference, and published labels from the other datasets were harmonized to these seven populations (**Supplementary Table 3**). Cell-type-specific accessible chromatin regions was defined using scATAC-seq resources from our normal-colon dataset^26^ and the Stanford HTAN colorectal atlas^17^, and merged for cell populations represented in both datasets (**Methods; Supplementary Table 4**). The resulting annotations contained 55,454-152,781 accessible regions across epithelial populations (**Extended Data Fig. 1c; Supplementary Table 4**) and were used to define the candidate regulatory variants for GReX prediction.

For colorectal polyps, we analyzed matched scRNA-seq and genotype data from 84 participants in the HTAN COLON MAP^24^ study. Nine epithelial populations were defined, including the seven presented in normal mucosa plus adenoma-specific cells (ASC) and serrated-polyp-specific cells (SSC) (**Extended Data Fig. 1d**). Accessible chromatin regions in polyp epithelial populations were defined by integrating our polyp scATAC-seq dataset with scATAC-seq data from 48 adenomas in the Stanford HTAN atlas (**Methods**). Across the nine populations, the resulting annotations comprised 13,061-188,833 accessible regions (**Extended Data Fig. 1e; Supplementary Table 4**).

Under our sTF-TWAS, we developed cell-type-specific GReX prediction models using matched single-cell RNA-seq and genotype data (**Methods; Extended Data Figs. 2** and 3). We integrated the cell-type-specific GReX models with summary statistics from two CRC GWAS meta-analyses: a European-ancestry analysis of 99,260 cases and 1,119,512 controls and a multi-ancestry analysis of 132,261 cases and 1,752,398 controls, which combined previously published CRC GWAS results^1^, with new data from BioVU^27^, the Million Veteran Program^28^, FinnGen release 13^29^, the UCLA ATLAS Community Health Initiative^30^, the Michigan Genomics Initiative Freeze 6^31^ and the Taiwan Precision Medicine Initiative^32^ (**Methods; Extended Data Figs. 2 and 3; Supplementary Table 5**). Significant associations from scTWAS analyses were further evaluated using Bayesian colocalization^33^ and summary-data-based Mendelian randomization with the heterogeneity in dependent instruments test (SMR-HEIDI)^34^. We then evaluated prioritized genes across complementary regulatory and functional evidence layers (**Fig. 1b-d**): cell-type-specific TF-target networks anchored on 51 previously identified CRC susceptibility-associated TFs^23^ (**Supplementary Table 6**); differential analyses using independent single-cell, spatial, TCGA-COAD^35^ and CPTAC-COAD^36^ datasets spanning normal mucosa to carcinoma, and assessed therapeutic relevance using DrugBank^37^, ChEMBL^38^, Therapeutic Target Database^39^, and Open Targets^40^, together with transcriptional signature-reversal analyses. Finally, we evaluated cancer dependency using DepMap^41^ and performed experimentally tested selected genes using CRISPR-Cas9 (**Fig. 1e**). Accessions, versions and access conditions of all resources are listed in **Supplementary Table 1**.

### Cell-type-specific susceptibility genes in normal colon epithelium cells

Across the model training in seven normal-colon populations in the two cohorts, 2,305-8,720 genes per cell type met prediction-performance criteria (cross-validated R² > 0.01) and were retained for downstream association analyses (**Methods; Extended Data Fig. 4a**). Association *P* values obtained independently from the two cohorts were combined with the Cauchy combination test (**Methods**). Integrating the models with the European-ancestry CRC GWAS meta-analysis, we identified 305 gene-cell-type associations at cell-type-specific Bonferroni-corrected *P* < 0.05: 69 in ABS, 42 in CT, 14 in EE, 66 in GOB, 53 in STM, 42 in TAC and 19 in TUF cells (**Fig. 2a-g; Supplementary Table 7**). Of these, 105 had additional evidence supporting a shared genetic signal between gene expression and CRC risk, defined by either strong colocalization (PP.H4 > 0.8) or significant SMR with no evidence of heterogeneity by HEIDI (**Methods; Supplementary Table 7**). Applying the same models to the multi-ancestry CRC GWAS identified 92 additional gene-cell-type associations beyond those detected in the European-ancestry analysis, comprising 24 in ABS, 14 in CT, 3 in EE, 16 in GOB, 20 in STM, 9 in TAC and 6 in TUF cells (**Extended Data Fig. 5; Supplementary Table 8**), 28 of which had colocalization or SMR-HEIDI support.

**Figure 2.**
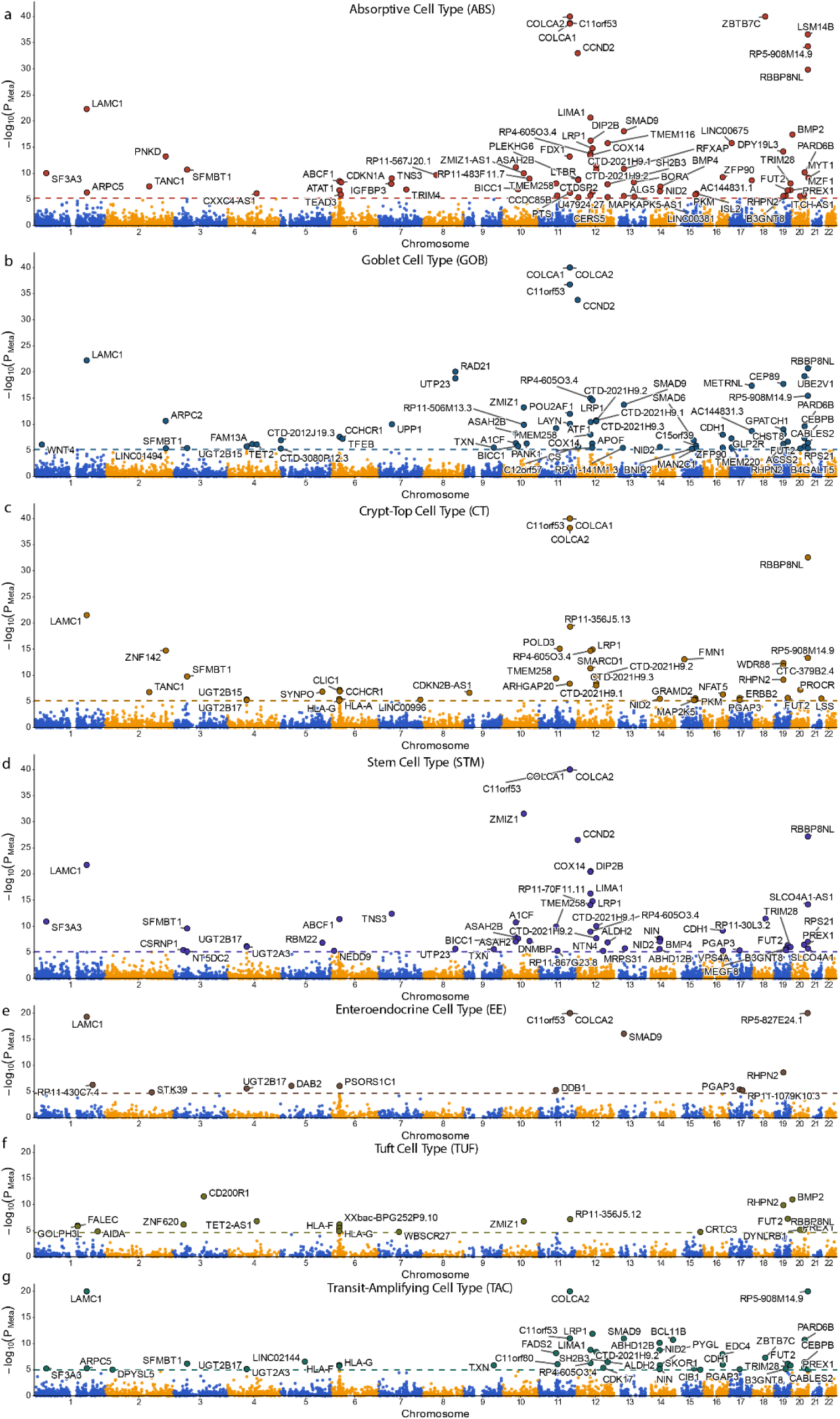
Genome-wide distribution of CRC risk genes identified by scTWAS in normal colon. **a–g**, Cell-type-specific scTWAS associations for absorptive (a), goblet (b), crypt-top (c), stem (d), enteroendocrine (e), tuft (f) and transit-amplifying (g) cells. Each point represents a gene, plotted by chromosomal position against −log10 of the Cauchy-combined *P* value across the two normal-colon cohorts. Alternating colors distinguish consecutive chromosomes. Dashed horizontal lines indicate the cell-type-specific Bonferroni significance thresholds. Enlarged and labelled points denote genes reaching significance in the corresponding cell type. Full association results are provided in Supplementary Table 7.

In total, the European-ancestry and multi-ancestry analyses identified 397 gene-cell-type associations involving 241 unique genes in normal colorectal epithelium (**Supplementary Tables 7 and 8**). Of these genes, 127 had not previously been implicated by CRC genetic studies^1–3,8–14^, substantially expanding the set of candidate susceptibility genes and resolving their genetically regulated effects across defined epithelial populations. The strongest associations were at 11q23.1, where *COLCA1* (EUR *P* = 9.3 × 10⁻⁵⁰, STM), *COLCA2* (*P* =2.4 × 10⁻⁴⁸) and *C11orf53* (*P* = 1.9 × 10⁻⁴⁷) were each significant in at least four epithelial populations with colocalization support. In total 35 genes were highlighted in a single cell population with colocalization or SMR-HEIDI support, 17 of them previously unreported, including *ZNF142* in crypt-top cells (*P* = 1.8 × 10⁻¹⁵, PP.H4 = 0.90) and *CD200R1* in tuft cells (*P* = 2.8 × 10⁻¹², SMR *P* = 9.9 × 10⁻⁴). These cell-type-restricted signals highlight susceptibility genes that may be obscured in bulk-tissue analyses.

### Cell-type-specific susceptibility genes in colorectal polyps cells

We next conducted scTWAS to colorectal polyps based on model training from 84 individuals. Across the nine cell-type-specific models, a median of 1,227 genes per cell type met the prediction-performance criteria (**Extended Data Fig. 4b**). Integrating with the European-ancestry CRC GWAS, we identified 109 gene-cell-type associations at Bonferroni-corrected *P* < 0.05: 21 in ASC, 10 in SSC, 31 in ABS, 23 in GOB, 9 in STM and 10 in TAC cells, with an additional 5 associations across EE, CT and TUF cells (**Fig. 3a-f; Supplementary Table 9**). Twenty-one associations were supported by colocalization or SMR-HEIDI, including 10 in ABS, 4 in GOB, 2 in STM, 2 in TAC, and 3 in other cell populations. The multi-ancestry analysis identified an additional 40 associations, six of which were supported by colocalization or SMR-HEIDI (**Extended Data Fig. 6; Supplementary Table 10**). The polyp models therefore yielded 149 associations involving 137 unique genes, 94 of which had not been implicated previously^1–3,8–14^.

**Figure 3.**
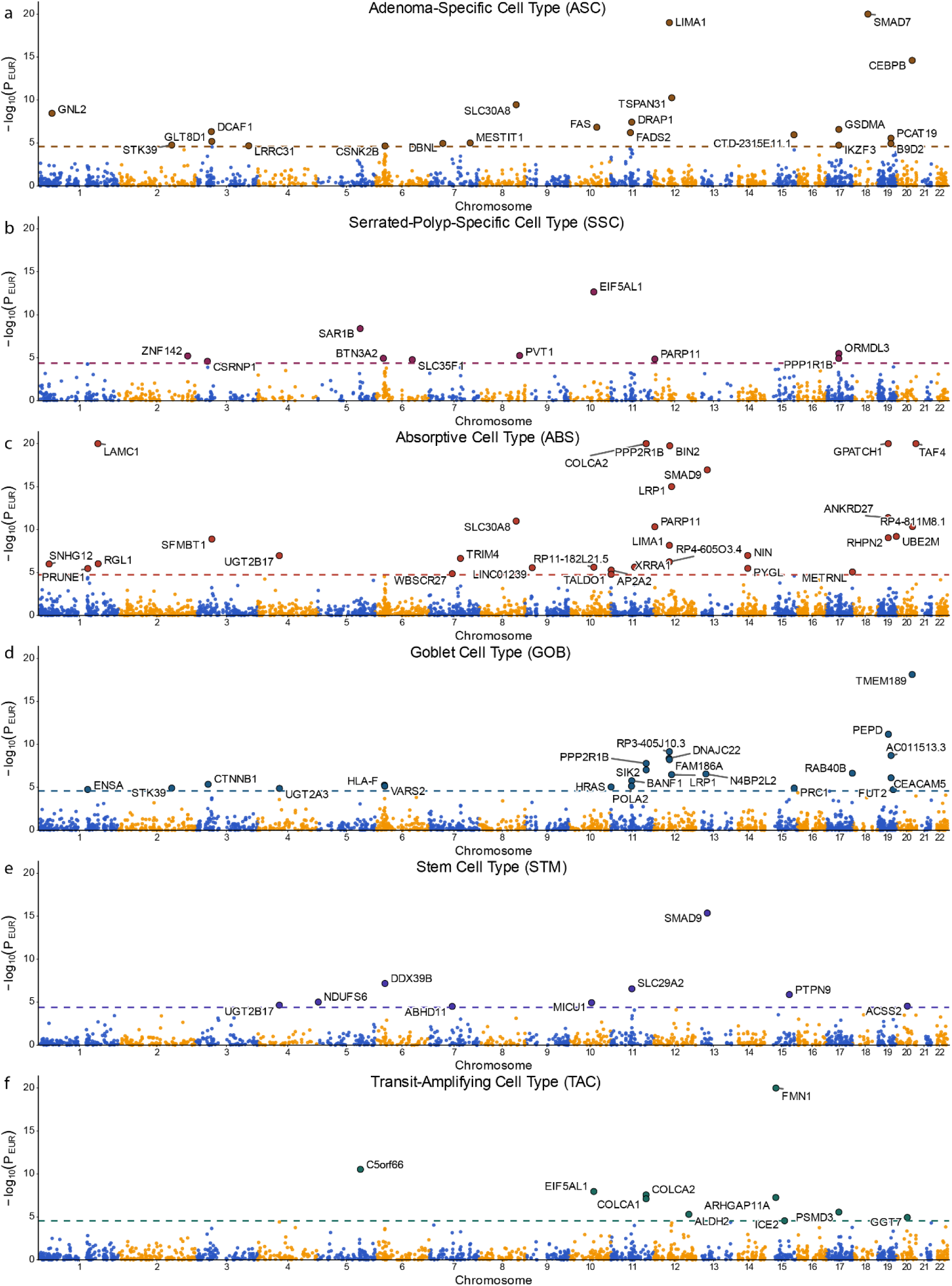
Genome-wide distribution of CRC risk genes identified by scTWAS in colorectal polyps. **a–f**, Cell-type-specific scTWAS associations for adenoma-specific (a), serrated-polyp-specific (b), absorptive (c), goblet (d), stem (e) and transit-amplifying (f) cells. Each point represents a gene, plotted by chromosomal position against −log10 of the European-ancestry scTWAS *P* value. Alternating colors distinguish consecutive chromosomes. Dashed horizontal lines indicate the cell-type-specific Bonferroni significance thresholds. Enlarged and labelled points denote genes reaching significance in the corresponding cell type. Full association results are provided in **Supplementary Table 9**.

Combining all 16 epithelial models, we identified 546 gene-cell-type associations involving 347 unique CRC susceptibility genes, 213 of which had not been implicated previously^1–3,8–14^. Among the seven cell populations represented in both tissue stages, 397 associations were found in normal mucosa and 102 in polyps, and only 17 associations involving 15 genes were shared (**Fig. 4a**). The lesion-restricted ASC and SSC models contributed a further 32 and 15 associations, respectively, with no gene shared between them. Overall, 258 genes (74%) were prioritized in only one epithelial population, indicating substantial cell-type specificity (**Fig. 4a; Supplementary Table 11**).

**Figure 4.**
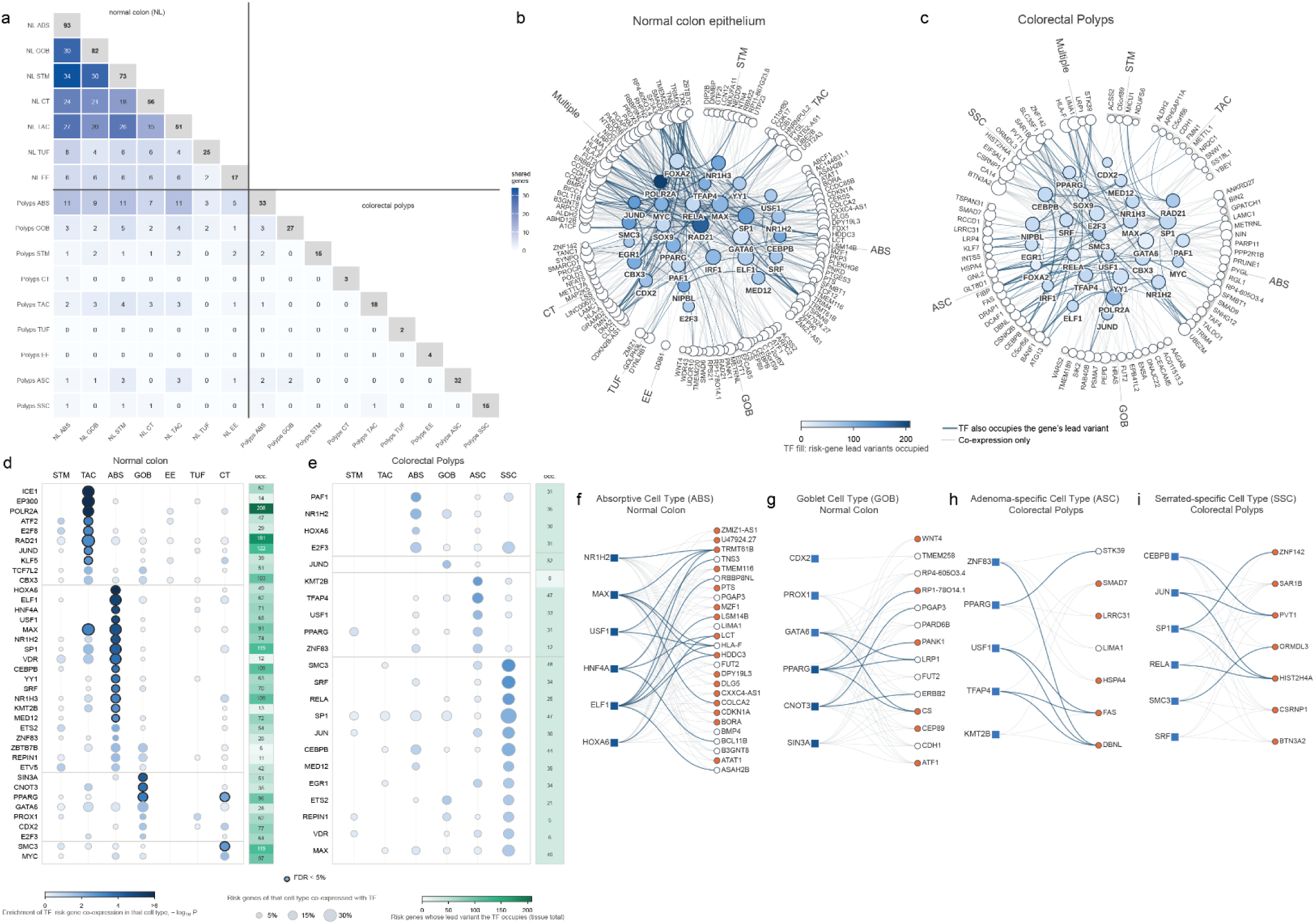
Susceptibility transcription factors organize cell-type-specific CRC risk-gene networks. **a)**, Overlap of risk genes across 16 epithelial scTWAS models. Diagonal cells indicate the number of genes prioritized in each model; off-diagonal cells indicate the number shared between pairs of models. **b),c),** TF–risk gene networks in normal colon (b) and colorectal polyps (c). Risk genes are grouped by the epithelial population in which they were prioritized. TF nodes are positioned centrally, with node fill indicating the number of risk-gene lead variants occupied by the TF. Edges denote TF–risk gene co-expression at Pearson *r* > 0.5; darker edges additionally indicate TF occupancy of the corresponding risk-gene lead variant. **d),e)**, Cell-type enrichment of TF–risk gene relationships in normal colon (d) and polyps (e). Dot size indicates the proportion of a cell type’s risk genes co-expressed with each TF, and color indicates enrichment relative to the remaining populations within the same tissue. Black outlines denote FDR < 0.05. **f)–i)**, Cell-type-specific TF–risk gene modules for absorptive (f) and goblet (g) cells in normal colon and ASC (g) and SSC (i) in polyps. Filled target nodes denote genes unique to that population; open nodes denote genes shared with at least one other model. Edges indicate *r* > 0.5, with darker edges denoting *r* > 0.7.

### Susceptibility TFs organize risk genes into cell-type-specific regulatory networks

To investigate whether susceptibility associations converge on shared transcriptional regulators, we analyzed 51 previously identified CRC susceptibility-associated TFs^14^, integrating TF-risk gene co-expression in donor-level pseudobulk profiles with TF ChIP-seq occupancy at GReX lead variants (**Methods**). Co-expression analyses identified 982 TF-gene pairs in normal mucosa and 415 in polyps (Pearson’s r > 0.5; FDR < 0.05), involving 131 and 95 susceptibility genes, respectively (**Supplementary Table 14; Extended Data Fig. 7a,c**). ChIP-seq analyses identified 226 GReX lead variants overlapping TF-binding peaks, providing occupancy support for 160 of 241 susceptibility genes in normal mucosa and 93 of 137 in polyps (**Supplementary Table 15; Extended Data Fig. 7b,d**). These variants overlapped binding peaks for a median of 11 and 8 TFs, respectively, with POLR2A, RAD21, JUND and SP1 among the most frequently represented. The two evidence layers were only weakly correlated across TFs within cell types (Spearman ρ = −0.21 to 0.37; **Extended Data Fig. 8a**), and 55 genes in normal mucosa and 37 in polyps were supported by both (**Extended Data Fig. 8b**).

Integrating both layers, we prioritized 28 TFs from the top 20 ranked in each tissue stage, 12 of which were shared (**Methods; Extended Data Fig. 8c,d**). These TFs formed candidate regulatory networks connecting 124 susceptibility genes through 579 TF-gene links in normal mucosa and 81 genes through 292 links in polyps (**Fig. 4b,c; Supplementary Tables 15 and 16**). In normal mucosa, GATA6, MAX, RAD21, SP1 and ELF1 were among the most prominent TFs, each associated with numerous susceptibility genes through co-expression and/or TF occupancy. GATA6 and MAX showed particularly extensive co-expression with susceptibility genes, whereas RAD21, POLR2A and SP1 exhibited broad occupancy at risk-associated variants. In polyps, SP1, RAD21 and POLR2A remained prominent, although their associated gene sets were generally smaller than those in normal mucosa. Other TFs, including SMC3, CBX3, CEBPB and TFAP4, showed evidence of regulatory associations in both tissue stages.

We next tested whether TF-risk gene relationships were enriched within specific epithelial populations (**Methods**). In normal mucosa, 28 TF-cell-type combinations were significantly enriched (FDR < 0.05), predominantly in ABS, TAC, GOB and CT cells (**Fig. 4d; Supplementary Table 17**). In polyps, enrichments within ASC and SSC reached only nominal significance (**Fig. 4e**). Representative networks illustrate these differences (**Fig. 4f-i**). In normal mucosa, ABS networks prominently featured HNF4A, MAX and HOXA6, and GOB networks featured GATA6, CDX2 and PPARG, with candidate susceptibility genes such as WNT4, CDH1 and ERBB2. In polyps, ASC networks were characterized by KMT2B and TFAP4, and SSC networks by CEBPB, JUN, RELA and SP1. These analyses reveal distinct TF-susceptibility gene network organization across epithelial populations and between normal and premalignant colorectal tissues.

### Independent multi-omic and spatial analyses support prioritized genes

We assessed whether scTWAS-prioritized genes showed concordant expression changes across tumorigenesis using donor-level pseudobulk differential-expression analyses of normal epithelium, adenoma, serrated polyp and carcinoma in the COLON MAP single-cell resource (**Methods**). Across eight epithelial-stage contrasts: normal mucosa versus ASC, SSC, microsatellite instability-high (MSI-H) or microsatellite-stable (MSS), and ASC or SSC versus MSI-H or MSS (**Fig. 5a**), we evaluated 397 normal-colon and 102 shared-polyp gene-cell-type associations. Along the direct normal-to-carcinoma trajectory, 118 (29.7%) normal-colon and 27 (26.5%) shared-polyp associations showed concordant expression changes in at least one MSI-H or MSS carcinoma comparison (**Fig. 5a; Supplementary Table 18**). Along the adenoma trajectory, 104 and 17 associations were supported in the normal-to-ASC transition and 87 and 20 in the ASC-to-carcinoma transition; corresponding numbers along the serrated trajectory were 108 and 29 (normal-to-SSC) and 123 and 31 (SSC-to-carcinoma). Among 32 ASC-specific genes, 12 and 11 showed concordant changes relative to normal STM and TAC cells, respectively, and two and four were additionally supported in ASC-to-MSI-H and ASC-to-MSS carcinoma comparisons. Among 15 SSC-specific genes, five showed concordant changes relative to normal GOB cells, and one and two were supported in SSC-to-MSI-H and SSC-to-MSS comparisons (**Fig. 5b; Supplementary Table 19**).

**Figure 5.**
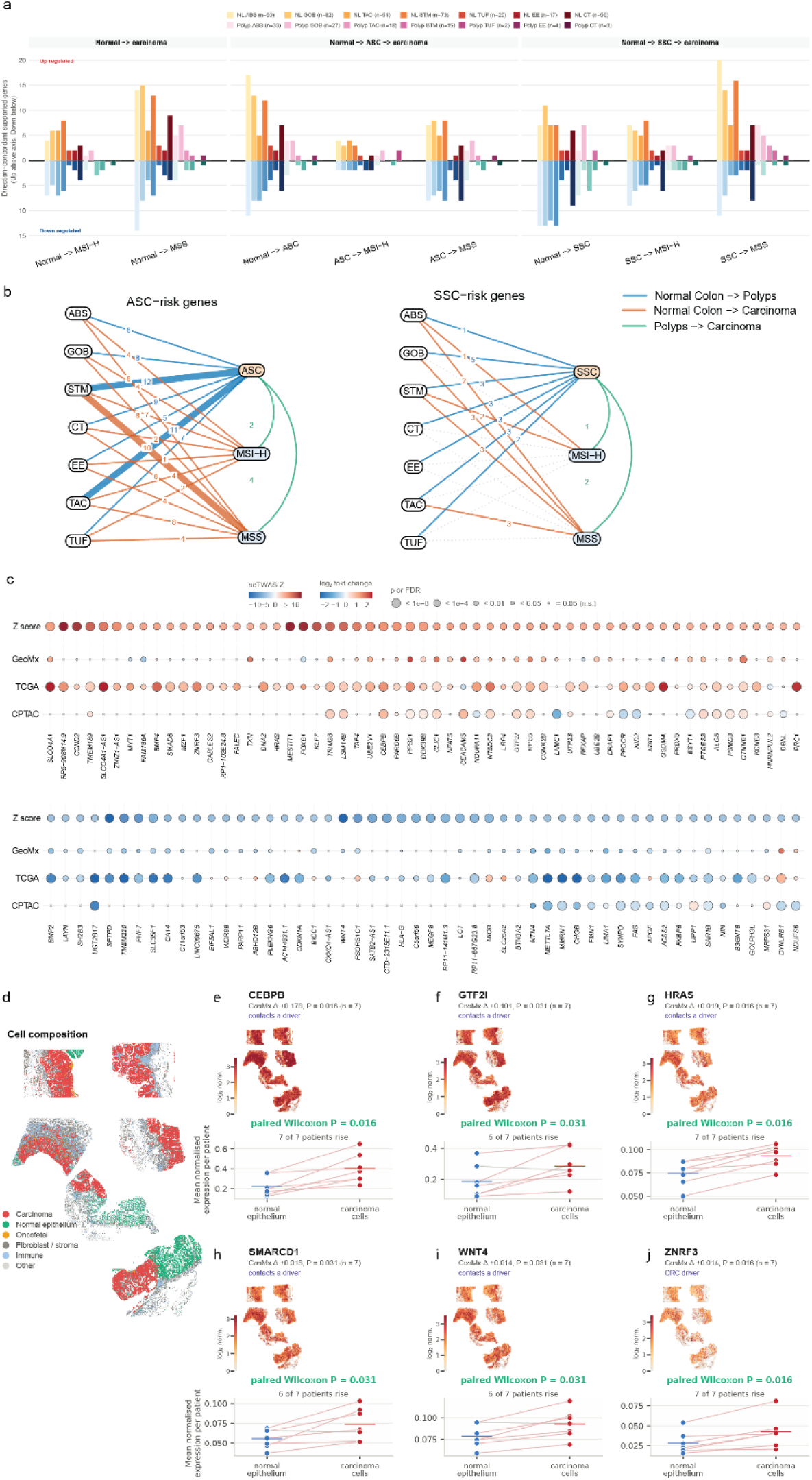
Orthogonal transcriptomic, proteomic and spatial support for scTWAS-prioritized CRC susceptibility genes. **a**, Cell-type-specific pseudobulk differential-expression support for scTWAS gene–cell-type associations across epithelial transitions from normal colon to MSI-H or MSS carcinoma, either directly or through adenoma-specific (ASC) or serrated-polyp-specific (SSC) states. Support required FDR < 0.05, |log2 fold change| > 0.5 and directional concordance with the scTWAS Z-score. Positive and negative bars indicate increased and decreased expression, respectively, in the later state. **b,** Differential-expression support for genes prioritized by the ASC and SSC scTWAS models across normal-to-polyp, normal-to-carcinoma and polyp-to-carcinoma transitions. Edge labels indicate the number of directionally concordant genes, with thicker edges denoting greater support; transitions without supported genes are omitted. **c,** Orthogonal tumor–normal support for scTWAS-prioritized genes across GeoMx PanCK-positive epithelial spatial transcriptomics, TCGA-COAD transcriptomics and CPTAC-COAD proteomics. Genes are grouped by representative scTWAS direction into positive (Z > 0) and negative (Z < 0) associations. Shown are genes with directionally concordant support from at least one resource. Bubble color represents effect size and bubble size represents statistical significance; absent bubbles indicate genes not measured in the corresponding dataset. **d,** CosMx single-molecule spatial profiling of early CRC, showing the spatial distribution of major cell-lineage populations. **e–j,** Spatial expression and paired carcinoma-versus-normal epithelial expression of *CEBPB, GTF2I, HRAS, SMARCD1, WNT4* and *ZNRF3*, selected according to prespecified criteria described in Methods. Paired comparisons are based on patient-level mean expression among patients with matched carcinoma and normal epithelial populations; *P* values were calculated using paired Wilcoxon signed-rank tests (Supplementary Table 21).

We then sought orthogonal support for all 347 genes in spatial GeoMx transcriptomic data^42^, TCGA-COAD^35^ transcriptomes and CPTAC-COAD^36^ proteomes (**Methods**). By the direction of the scTWAS association, 176 genes were risk-increasing and 171 risk-decreasing. Overall, 109 genes showed directionally concordant evidence in at least one of the three datasets, including 57 risk-increasing and 52 risk-decreasing genes. Among the risk-increasing genes, 39 were supported by one dataset, 17 by two datasets and one by all three. The corresponding numbers for risk-decreasing genes were 32, 16 and four, respectively. Genes not measured on a platform were treated as untested rather than unsupported (**Fig 5c; Supplementary Table 20**). Combing all the omics-analysis, 233 genes (67%) had at least one directionally concordant differential-expression result across the single-cell, spatial, transcriptomic and proteomic datasets (**Supplementary Table 20**).

In addition, we examined the spatial localization of prioritized genes using CosMx single-molecule imaging of 11 early CRCs, comprising 1,257,627 quality-controlled cells (**Fig. 5d**). Seven tumors contained both carcinoma and residual normal or undefined epithelial compartments and were used for paired within-patient comparisons. Of the 233 susceptibility genes supported by the above analysis, 91 were represented on the CosMx panel, of which 27 showed nominally significant differential expression between the two compartments (paired Wilcoxon signed-rank *P* < 0.05), 20 of them in association directions consistent with the scTWAS (**Supplementary Table 21**). Among these, several established CRC-related genes, including *CEBPB, GTF2I, HRAS, WNT4, ZNRF3* and *SMARCD1*, exhibited higher expression in carcinoma cells in six or seven of the seven patients analyzed (**Fig. 5e-j**).

### Druggable susceptibility genes identify candidate therapeutic vulnerabilities

To assess the therapeutic relevance, we integrated drug-target relationships from DrugBank^37^, ChEMBL^38^, Therapeutic Target Database^39^, and Open Targets^40^. Of the 347 susceptibility genes, 63 (18.2%) encoded druggable proteins linked to 303 approved or clinical-stage compounds (**Supplementary Table 22**). Mechanism-of-action information was available for 172 drug-target interactions involving 43 genes and was predominantly inhibitory. Restricting the analysis to approved or phase II/III compounds identified 55 susceptibility genes linked to 193 clinically advanced drugs (**Fig. 6**). This recovered established therapeutic relationships, including ERBB2, targeted by trastuzumab, pertuzumab, tucatinib, neratinib, lapatinib and margetuximab, and identified additional genetically supported drug-target pairs: the proteasome components PSMA7, PSMD3 and PSMD13 (bortezomib, carfilzomib and ixazomib), DDR1 (imatinib), POLD3 (gemcitabine and cytarabine) and the mitochondrial complex I components NDUFA11 and NDUFS6 (metformin). Twenty-nine of the 347 susceptibility genes were represented in the COSMIC Cancer Gene Census^43^, with enrichment among druggable genes (11 of 63 versus 18 of 284; odds ratio = 3.1, two-sided Fisher’s exact P = 0.009). KEGG Pathway enrichment analysis placed the druggable susceptibility genes within signaling, proteostasis and metabolic programs relevant to CRC biology and treatment (**Methods, Supplementary Table 23**).

**Figure 6.**
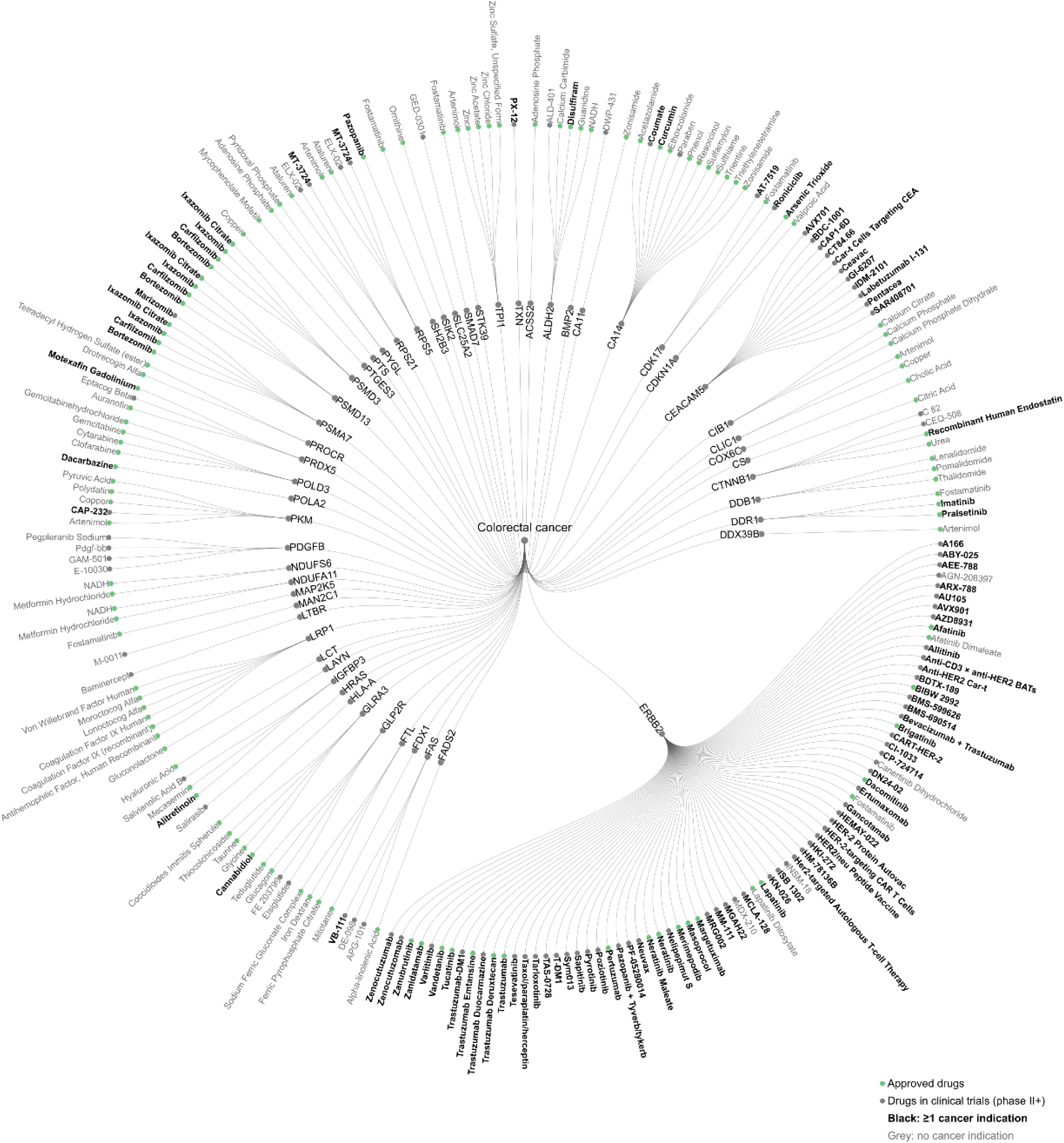
Therapeutic annotation of cell-type-specific CRC susceptibility genes. Druggable CRC risk genes are shown as inner nodes, with approved or clinical-stage compounds annotated to each gene shown as outer nodes. Drug–gene relationships were compiled from DrugBank, ChEMBL, the Therapeutic Target Database and Open Targets (Supplementary Table 22). Green markers denote approved drugs, whereas grey markers denote compounds in phase II or later clinical trials. Compound names shown in black have at least one cancer indication, whereas names shown in grey have no annotated cancer indication.

We next tested whether approved drugs targeting susceptibility genes have potential to reverse risk-associated cell-type-specific transcriptional signatures. Among 43 compounds assessed using the iLINCS^44^ large-intestine perturbation dataset, four showed significant negative connectivity after Benjamini-Hochberg correction: the proteasome inhibitors ixazomib (normalized connectivity score (NCS) = −0.19, FDR = 0.0043), targets PSMA7, PSMD3 and PSMD13, the *ERBB2* inhibitor neratinib (NCS = −0.17, FDR = 0.014), bortezomib (targets *PSMA7, PSMD3 and PSMD13,* NCS = −0.15, FDR = 0.0043) and resorcinol (target *CA14,* NCS = −0.63, FDR = 0.019). These findings provide transcriptional perturbation-based support for prioritizing candidate drugs for further investigation in CRC prevention (**Methods; Extended Data Fig. 9; Supplementary Table 24**).

### CRC susceptibility genes are enriched for cancer-cell dependencies and validated by genetic perturbation

We next evaluated whether the identified susceptibility genes contribute to CRC cell fitness. We compared median DepMap^41^ gene-effect scores for 274 evaluable scTWAS risk genes with those for 466 background genes, 175 CRC consensus susceptibility genes and 52 Tier 1 colorectal driver genes of the COSMIC Cancer Gene Census (**Methods; Supplementary Table 25**). scTWAS risk genes showed stronger dependency than background genes (median gene effect, −0.078 versus −0.018; Holm-adjusted *P* = 1.1 × 10⁻⁶) and CRC consensus genes (median, −0.037; *P* = 0.043). Their dependency profile was comparable to that of established CRC drivers (median, −0.083; 17.3% with gene-effect scores < −0.5 versus 13.1% of risk genes; *P* = 0.71) (**Fig. 7a**).

**Figure 7.**
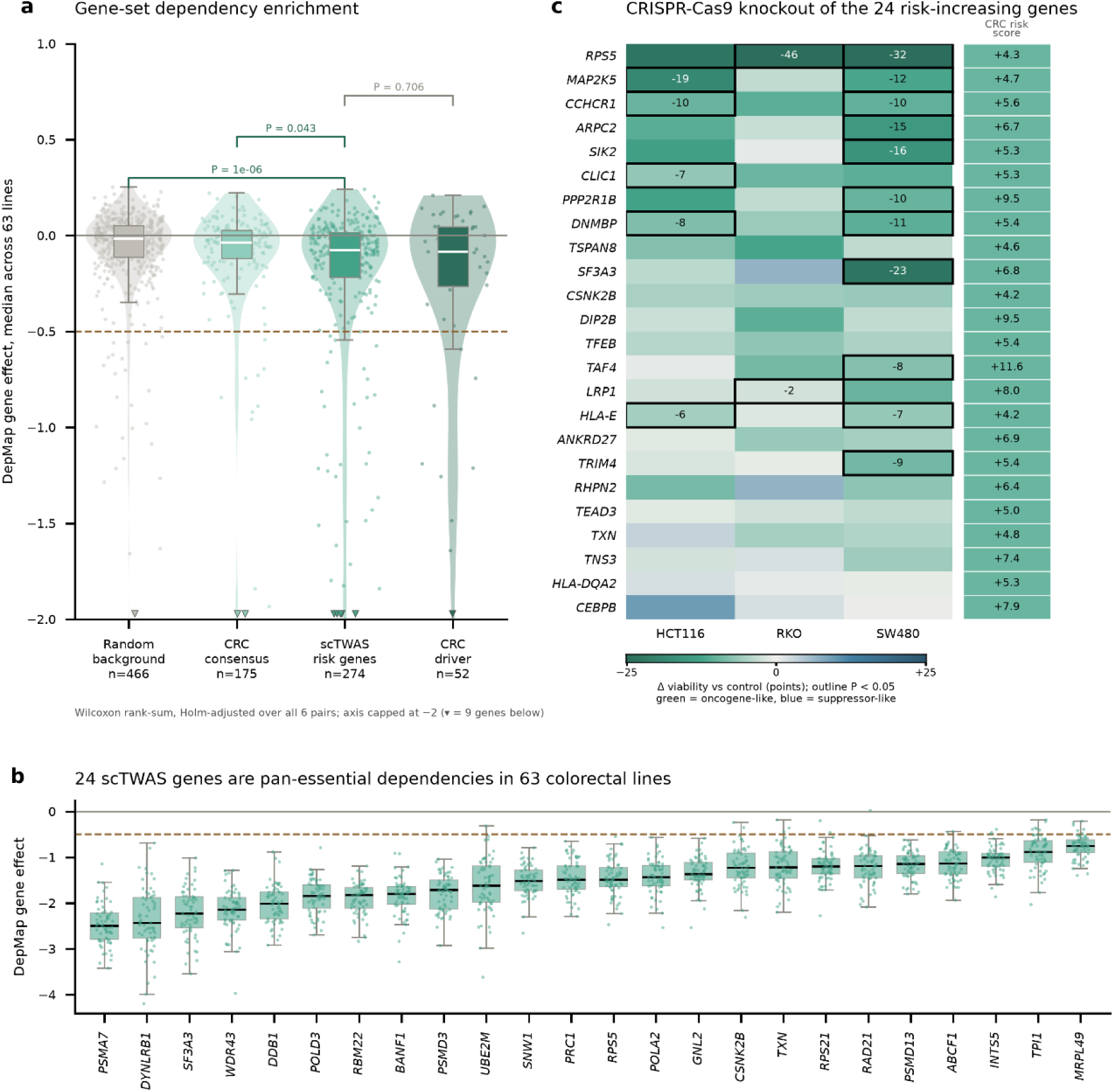
Functional dependency and experimental validation of scTWAS-prioritized CRC risk genes. **a**, DepMap CRISPR gene-effect distributions across four gene sets: a random background set, the CRC consensus set, scTWAS-prioritized risk genes and established CRC somatic drivers. Only genes scored in more than 30 colorectal adenocarcinoma cell lines were included. Violins show gene-level distributions, boxes indicate the interquartile range and median, and points represent individual genes. Pairwise differences were assessed using Wilcoxon rank-sum tests with Holm correction across all six comparisons; brackets indicate adjusted *P* values. The dashed horizontal line marks the −0.5 dependency threshold. **b,** DepMap CRISPR gene effects for the 24 scTWAS genes classified as pan-essential dependencies across 63 colorectal adenocarcinoma cell lines, defined by a median gene effect below −0.5, measurements in more than 30 lines and gene effects below −0.5 in at least 90% of scored lines. Genes are ordered from the strongest to weakest median dependency. Boxes indicate the interquartile range and median, whiskers extend to 1.5× the interquartile range, and points represent individual cell lines. The dashed horizontal line marks the −0.5 dependency threshold. **c,** Arrayed CRISPR–Cas9 knockout of 24 risk-increasing scTWAS genes in HCT116, RKO and SW480 CRC cell lines. Heatmap colors indicate the change in cell viability relative to same-plate non-targeting controls, with negative values indicating reduced viability after knockout. Outlined cells denote nominal significance by paired *t*-test (*P* < 0.05), with effect sizes shown within significant cells. The adjacent strip shows the corresponding CRC scTWAS Z-score for each gene. All three cell lines passed assay plate-quality-control criteria.

Twenty-four prioritized genes had gene effects below −0.5 in at least 90% of colorectal adenocarcinoma cell lines (**Methods; Fig. 7b; Supplementary Table 25**). These included components of the proteasome (*PSMA7, PSMD3* and *PSMD13*), RNA processing and splicing machinery (SF3A3, RBM22 and SNW1), ribosomal and translational machinery (*RPS5, RPS21* and *MRPL49*), and DNA replication, repair and chromosome organization (*DDB1, POLD3, POLA2, RAD21* and *BANF1*), indicating that a subset of inherited susceptibility genes maps to cellular processes broadly required for CRC-cell fitness. To evaluate risk-increasing genes directly, we additionally performed arrayed CRISPR-Cas9 knockout of 24 selected scTWAS genes in HCT116, RKO and SW480 CRC cells (**Methods; Fig. 7c; Supplementary Table 26**). Thirteen genes significantly reduced cell viability in at least one cell line at nominal two-sided P < 0.05, yielding 18 significant gene-cell-line effects. All 18 were in the direction expected for genes whose increased genetically regulated expression is associated with higher CRC risk (one-sided binomial *P* = 3.8 × 10⁻⁶). Eleven significant effects were observed in SW480, five in HCT116 and two in RKO cells. The largest effects were for *RPS5* (−46 percentage points in RKO and −32 in SW480), *SF3A3* (−23 in SW480) and *MAP2K5* (−19 in HCT116), and five genes (*RPS5, MAP2K5, CCHCR1, DNMBP* and *HLA-E*) had significant effects in more than one cell line. Together, these analyses identified 35 distinct scTWAS-prioritized genes with evidence of cellular fitness effects, comprising 24 DepMap dependencies and 13 arrayed CRISPR hits, with two genes *(RPS5* and *SF3A3*) supported by both approaches.

## Discussion

We present a cell-type-resolved map of inherited CRC susceptibility, built by learning GReX from matched genotype and scRNA-seq data in normal colorectal epithelium and premalignant polyps. Our previous studie^14,22,23^, have demonstrated through simulation and real-data analyses that incorporating regulatory annotations into variant selection can improve TWAS performance and risk-gene discovery. Building on this principle, we restricted predictors to putative regulatory variants within cell-type-specific accessible chromatin regions and tested 16 epithelial models against European-ancestry and multi-ancestry GWAS meta-analyses of up to 132,261 cases. We identified 546 gene-cell-type associations implicating 347 genes, 213 of which had not been linked to CRC susceptibility by earlier CRC genetic studies^1–3,8–14^. A substantial fraction of associations had additional support from colocalization or SMR-HEIDI. Our findings extend conventional bulk-tissue TWAS by resolving susceptibility associations across distinct epithelial populations and premalignant states, providing new insights into the cellular contexts and regulatory mechanisms through which inherited genetic variation may contribute to CRC development.

Our findings further reveal that inherited CRC susceptibility is associated with distinct epithelial cell populations and premalignant states rather than uniformly distributed across the colorectal epithelium. Notably, 74% of the 347 prioritized genes were identified in a single cell-type model. Adenoma-associated and serrated-polyp-associated epithelial populations contributed 47 associations (32 and 15, respectively), with no genes shared between these populations and 36 genes uniquely prioritized in these premalignant states. Particularly, among the 32 ASC-specific genes, 12 and 11 showed differential expression between STM and ASC and between TUF and ASC, respectively, whereas five of the 15 SSC-specific genes were differentially expressed between GOB and SSC. These findings highlight the importance of cell-type-resolved analyses in uncovering genetic regulatory effects that may be diluted or obscured in bulk tissue, while identifying the epithelial contexts in which inherited susceptibility may operate. Importantly, the identification of susceptibility genes in premalignant epithelial populations extends previous TWAS beyond normal colorectal tissue and provides insights into potentially distinct genetic regulatory mechanisms underlying conventional adenoma and serrated pathways. Nevertheless, these lesion-associated findings require validation in independent polyp cohorts to establish their reproducibility and biological relevance.

The TF-network analysis adds regulatory organization beyond gene lists. Lineage-defining factors including GATA6, MAX and RAD21 linked to many risk genes in normal mucosa, with HNF4A and HOXA6 featuring in absorptive and CDX2, GATA6 and PPARG in goblet networks, whereas SP1 was the most connected factor in polyps; yet only 13 of 871 TF-gene links were shared between tissue stages. In polyps, TFAP4 and KMT2B characterized adenoma-specific and CEBPB, JUN, RELA and SP1 serrated-polyp-specific networks, pointing to distinct candidate regulatory programs in the two neoplastic routes. These enrichments reached only nominal significance, co-expression does not establish direct regulation, and the weak correlation between co-expression and occupancy indicates that the two layers capture different information. Moreover, because the tissue stages were profiled in different donor sets, limited sharing may partly reflect sampling and unequal power rather than tissue-stage-specific regulation. These networks are therefore hypotheses for perturbation in primary epithelial models.

Integration of independent multi-omics datasets provided complementary biological evidence supporting the prioritized CRC susceptibility genes. Overall, 233 genes (67%) exhibited directionally concordant differential expression in at least one single-cell, spatial transcriptomic, bulk transcriptomic or proteomic analysis, including 109 genes (31%) supported by GeoMx, TCGA or CPTAC datasets. These findings strengthen the biological relevance of genetically prioritized genes across colorectal epithelial states and tumor tissues, while providing insights into their potential involvement in colorectal tumorigenesis. Importantly, transcriptional and proteomic alterations in tumors may not consistently reflect the direction of genetically regulated expression in normal or premalignant epithelial cells, owing to somatic alterations, clonal selection and tumor microenvironmental remodeling. Thus, the absence of concordant differential expression does not necessarily weaken the underlying genetic associations. Rather, these orthogonal findings complement TWAS, colocalization and SMR–HEIDI evidence, collectively strengthening candidate gene prioritization and providing a foundation for subsequent functional investigation.

Integrating genetic evidence with druggability and functional perturbation analyses provides opportunities to prioritize therapeutic targets for CRC prevention. We identified 63 potentially druggable susceptibility genes (18.2%), including 55 targeted by 193 approved or late-stage compounds, with significant enrichment for known cancer genes (odds ratio = 3.1). Prioritized targets included ERBB2, proteasome components (PSMA7, PSMD3 and PSMD13), DDR1, POLD3 and mitochondrial complex I genes. Particularly, four (FADS2, FAS, SMAD7 and STK39) from ASC and one (CA14) from SSC. Notably, *SMAD7* has been implicated in CRC susceptibility through previous genetic studies^45,46^, whereas *FADS2* has been linked experimentally to lipid metabolism and ferroptosis sensitivity in CRC cells^47,48^. The identification of these genes in ASC and SSC models suggests that inherited CRC susceptibility may involve potentially actionable regulatory mechanisms in distinct premalignant epithelial states.Furthermore, transcriptional perturbation analyses identified significant reversal of CRC risk-associated expression signatures by ixazomib, bortezomib and neratinib, providing additional support for drug-repurposing opportunities. However, these findings require further evaluation of target directionality, tissue specificity and therapeutic safety, particularly because perturbations in established cancer cells may not reflect early tumorigenesis. Functional validation in primary adenoma organoids and *in vivo* chemoprevention models will be essential to establish the preventive potential of prioritized candidates.

Dependency analyses showed that scTWAS genes were more essential in CRC cell lines than background (median gene effect −0.078 versus −0.018) and consensus susceptibility genes, and comparable to established drivers, with 24 broadly shared dependencies spanning the proteasome, splicing, translation and DNA replication. Thirteen of 24 genes tested by arrayed CRISPR-Cas9 knockout reduced viability in at least one line, and all 18 significant effects were in the expected direction, giving 35 genes with fitness evidence in total. Genes such as *RPS5* and *SF3A3* play essential roles in fundamental cellular processes, including protein synthesis and RNA splicing, respectively, and their functions are not specific to colorectal biology. In contrast, *DNMBP* and *CLIC1*, exhibited concordant perturbation without widespread dependency across CRC cell lines, suggesting potentially more context-specific roles. These screens used nominal significance in three established malignant lines and do not test dependency in normal or premalignant epithelium.

Our study has several limitations. First, the performance of cell-type-specific GReX models depends on donor sample size, cell abundance and genetic regulatory effects. Rare epithelial populations and premalignant polyp models were particularly underpowered, as the latter were trained using only 84 donors compared with 381 for normal mucosa, yielding substantially fewer predictable genes. Consequently, fewer associations in polyps may reflect limited statistical power rather than fewer susceptibility mechanisms. Moreover, restricting genetic predictors to accessible chromatin regions may overlook distal regulatory variants or regulatory elements not captured by available scATAC-seq datasets. Larger cohorts with matched genotypes, single-cell transcriptomes and chromatin accessibility will improve model performance and expand gene discovery. Second, TWAS associations do not necessarily establish causal relationships, as linkage disequilibrium, correlated expression predictors and shared regulatory variants may implicate multiple genes at the same locus. Although colocalization and SMR–HEIDI analyses provided additional genetic support, they cannot fully exclude pleiotropy or complex allelic architectures. Furthermore, associations detected in only one cell type do not necessarily demonstrate exclusive cell-type-specific effects, particularly when statistical power differs across populations. Third, the predominance of European-ancestry genetic data and limited diversity of single-cell reference cohorts may restrict the generalizability of our findings across ancestral populations. Differences in allele frequencies, linkage disequilibrium and regulatory architectures warrant further evaluation in diverse populations. In addition, the relatively small polyp cohort may not fully capture the molecular heterogeneity of conventional adenomas and serrated lesions. Finally, although integrating TF–gene co-expression with ChIP-seq occupancy revealed candidate regulatory networks, these associations do not establish direct TF regulation or functional effects of susceptibility variants. Similarly, multi-omics evidence supports biological relevance but does not independently demonstrate causality. DepMap and CRISPR–Cas9 analyses identified genes involved in CRC cell viability, whereas drug-target annotations and transcriptional signature reversal highlighted potential therapeutic candidates; however, these findings may not directly translate to tumor initiation or prevention. Further validation using allele-specific assays, CRISPR perturbations, patient-derived adenoma organoids and in vivo models will be essential to establish regulatory mechanisms and evaluate preventive potential.

## Methods

### Ethics and study oversight

This study complied with all relevant ethical regulations. Human participants were enrolled under protocols approved by the responsible institutional or national ethics committees, with informed consent obtained as required. Secondary datasets were accessed and analyzed in accordance with applicable data-use agreements and provider terms. Participants in the VUMC HTAN COLON MAP were enrolled under protocols approved by the Vanderbilt University Medical Center (VUMC) Institutional Review Board (IRB #031078) and all participants provided written informed consent^16^. Participants in the VUMC Gut Cell Atlas were enrolled under a VUMC IRB-approved protocol (IRB #191738) with written informed consent^25^. Germline genotyping and single-cell transcriptomic profiling were conducted under the same IRB approvals. BioVU analyzes used de-identified whole genome sequencing (WGS) linked to electronic health records in the Vanderbilt Synthetic Derivative. The study was conducted under a VUMC IRB-approved protocol (IRB #250819) in accordance with BioVU data-use policies ^27^.

The Wellcome Sanger IBDverse study was approved by the NHS Research Ethics Committee, Cambridge South (REC17/EE/0338), and all participants provided written informed consent^21^. Single-cell and genotype data (EGAD00001015692 and EGAD00001016069) were accessed through the European Genome-phenome Archive following Data Access Committee approval.

Ethics approval and informed consent for external GWAS cohorts were obtained by contributing studies as described in the original publications. Only summary-level association data were analyzed here. Publicly available TCGA-COAD, CPTAC-COAD, GeoMx, CosMx, Stanford HTAN scATAC-seq, DepMap, Cistrome, iLINCS and drug-target resources were used in accordance with their respective access policies and terms of use. COSMIC Cancer Gene Census data were used under an academic license. Cell-line experiments using HCT116, RKO and SW480 did not involve newly recruited human participants.

### Cohorts and single-cell RNA sequencing

Normal-colon scRNA-seq data originated from 52 participants from VUMC HTAN COLON MAP and 51 from the VUMC Gut Cell Atlas, yielding 103 VUMC donors^15,23^. Sample collection, library preparation, sequencing, alignment and initial quality control have been described previously^15,23^. Briefly, both datasets were generated using a modified inDrop protocol^49^ with dual-indexed TruDrop libraries^50^, sequenced on an Illumina NovaSeq 6000, aligned to GRCh38 using STAR^51^ and processed into unique molecular identifier (UMI) counts using dropEst^52^. RNA-seq data in normal colon tissues from Gut Cell Atlas project were derived from 18 healthy control and 34 IBD patients with a prior history of CD who had normal histology on the index colonoscopy as “normals”. We preferentially retained ascending-colon samples from donors with multiple sampled sites, whereas COLON MAP data were restricted to normal mucosa. Genes detected in fewer than three cells and cells expressing fewer than 200 genes were excluded.

To harmonize cell-type annotations, we adopted seven COLON MAP epithelial populations as the reference vocabulary: ABS, CT, EE, GOB, STM, TAC and TUF. Fourteen cell type categories from Gut Cell Atlas annotation were mapped to the corresponding COLON MAP populations (**Supplementary Table 3**). Non-epithelial cells and ASC or SSC populations were excluded before integration. We performed single-cell normalization, integration, clustering and visualization were performed in Seurat v5.5.1^53^ under R v4.5.0. Each dataset was normalized separately using SCTransform^54^ (v0.4.3), regressing out mitochondrial transcript fraction. We selected 3,000 integration features, performed principal-component analysis and corrected for batch effects across three sequencing batches (one Gut Cell Atlas and two COLON MAP) using Harmony^55^ (v2.0.5). The first 30 Harmony dimensions were used for shared nearest-neighbor graph construction, Louvain clustering (resolution 0.8) and UMAP visualization^56^. Initial clustering identified 25 clusters, including 18 absorptive clusters and one or two clusters for each remaining epithelial population. We excluded eight clusters because their cell-type annotations were inconsistent with the expected epithelial populations, as assessed by cluster composition, UMAP distribution and canonical marker-gene expression. The retained cells were reprocessed from raw counts using the same normalization, integration and clustering workflow, yielding 120,891 cells across 25 clusters. Clusters were assigned their predominant harmonized cell-type label when it accounted for at least 60% of annotated cells. Twenty-three clusters met this threshold (minimum proportion, 71.9%); the remaining two (52.4% and 41.0%) were annotated based on cluster composition and canonical marker-gene expression. The final annotations resolved seven epithelial populations (ABS, CT, EE, GOB, STM, TAC and TUF) for downstream pseudobulk aggregation of expression for model building.

The Wellcome Sanger Institute IBDverse cohort contributed 278 normal-colon donors^21^. Sample collection, library preparation, sequencing, alignment, quality control and cell-type annotation have been described previously^21^. Briefly, libraries were generated using 10x Genomics Chromium 3′ v3.0/v3.1 chemistry, aligned to GRCh38 with Cell Ranger^57^ v7.2.0, and processed with CellBender^57^ v2.1 and Scrublet^58^ v0.2.1 to remove ambient RNA and multiplets. Published annotations were harmonized to the epithelial cell types used in this study (**Supplementary Table 3**). Moreover, polyps scRNA-seq data from 84 participants in the VUMC HTAN COLON MAP^15^, were processed following the published COLON MAP single-cell workflow^15^. For each donor and cell type, raw UMI counts were aggregated into pseudobulk expression count profiles.

### Genotyping, imputation and quality control

Germline genotype data were obtained from two sources: previously generated genotypes from the IBDverse cohort and newly generated genotypes from the VUMC HTAN Colon Map cohort. All genotype data were aligned to the GRCh38 human reference genome.

#### Previously generated genotype data from the IBDverse cohort

Genotype data for 278 participants were obtained from the European Genome-phenome Archive (EGA; accession EGAD00001016069). As described in the original study, DNA extracted from blood or gastrointestinal tissue was genotyped using the Applied Biosystems UK Biobank Axiom Array. Genotypes were jointly called across five batches and subsequently imputed using the Michigan Imputation Server^60^ with the TOPMed reference panel.

#### Newly generated genotype data from the VUMC HTAN Colon Map cohort

Germline genotyping was performed for 136 participants, including 52 with normal colorectal mucosa and 84 with colorectal polyps, using the expanded Illumina Multi-Ethnic Genotyping Array (MEGA). Initial variants were excluded for call rate below 95%, Hardy-Weinberg equilibrium P < 1 x 10^-6^ within genetic-ancestry groups, concordance below 92% across duplicate and 1000 Genomes Project quality-control samples, or minor allele frequency (MAF) below 0.5%. Samples were excluded for call rate below 95% or relatedness PI_HAT above 0.2, retaining one individual from each related pair. Genetic ancestry was inferred by principal-component analysis against 1000 Genomes Project reference populations and genetic sex was estimated with PLINK v1.9. Alleles were aligned to the forward strand with McCarthy Group Tools, variants with MAF below 1% were removed, and imputation was performed on the TOPMed Imputation Server against the TOPMed v3 reference panel^59^. Approximately 11 million variants passing the imputation-quality filter were retained for both cohorts. Following genotype quality control, approximately 1.8 million genotyped variants on GRCh38 were submitted to the TOPMed Imputation Server. Genotypes were phased using Eagle v2.4 and imputed against the TOPMed version R3 multi-ancestry reference panel using the server implementation of Minimac4^59,60^. After excluding variants with minor allele frequency <1% or imputation quality (*R*^2^ < 0.3), approximately 10.2 million variants were retained for downstream association analyses. Imputed allele dosages were used in the GWAS.

### Single-cell ATAC sequencing and cell-type peak definition

Cell-type-specific chromatin accessibility was derived from two independent resources. For the Stanford HTAN colorectal atlas^17^ (GSE201336), fragment files from 89 libraries were matched to the published sample metadata and analyses were restricted to normal-colon and colorectal-polyp samples. Barcode-level epithelial annotations released by the authors were harmonized to six populations (ABS, CT, EE, GOB, STM and TAC; **Supplementary Table 3**). Arrow files were generated against hg38 with ArchR^61^, retaining nuclei with transcription-start-site enrichment of at least 6 and at least 1,000 unique fragments. Peak-calling groups were defined by tissue condition and cell type, groups with fewer than 100 nuclei were excluded, and reproducible peaks were called from pseudobulk replicates with MACS2^62^, retained when supported by at least two replicates, capped at 150,000 peaks per group and filtered against the ENCODE hg38 blacklist v2^63^.

For the VUMC scATAC-seq resource^64^, pseudobulk BAM files were stratified by tissue condition and cell type, covering ABS, CT, EE, GOB, STM, TAC and TUF together with ASC and SSC. Alignments were restricted to properly paired reads with mapping quality of at least 30 on chromosomes 1-22 and X after removal of unmapped, secondary, duplicate, supplementary and quality-failed reads. Peaks were called per condition and cell type with MACS3 at q <= 0.01 and filtered against the ENCODE hg38 blacklist v2. For populations with fewer than one million usable fragments, reproducibility was assessed across three independent 80% subsamples and peaks supported in at least two of three subsamples were retained. Adenoma and serrated-polyp peaks were merged within each shared epithelial cell type, whereas ASC and SSC were retained as separate lesion-specific annotations.

For the six epithelial populations represented in both resources, accessible regions were integrated separately for normal colon and for polyps by combining the corresponding VUMC and Stanford HTAN peak sets, merging overlapping or directly adjacent intervals with bedtools merged 0^65^. TUF, ASC and SSC annotations were taken from VUMC resource alone. The resulting cell-type-specific accessible regions defined the candidate predictor space for GReX model construction.

### CRC GWAS meta-analysis

#### BioVU CRC case-control ascertainment

We performed a CRC GWAS in BioVU, the VUMC DNA biobank linked to de-identified electronic health records^27,66^. CRC cases were identified using ICD-9-CM codes 153 and 154 or ICD-10-CM codes C18-C20. To enrich for sporadic CRC, we excluded individuals with a documented family history of CRC (ICD-9-CM V16.0; ICD-10-CM Z80.0), inflammatory bowel disease (V18.59; Z83.79), or colorectal polyps (V18.51; Z83.71); those with familial adenomatous polyposis or Lynch syndrome; and those diagnosed with another malignancy (ICD-9-CM 140-209; ICD-10-CM C00-C96) more than six months before CRC diagnosis. Controls were defined as individuals without a recorded history of cancer. Genome-wide genetic variants were obtained from whole-genome sequencing (WGS) of DNA samples collected through Vanderbilt University Medical Center’s BioVU biobank^27,66^. Sequencing was performed on the Illumina NovaSeq 6000 platform with paired-end 150-bp reads, and variants were identified using the DRAGEN germline pipeline^67^ (v3.7.8) against the GRCh38 reference genome. BioVU DNA was extracted from residual blood specimens collected during routine clinical care. Sequence reads were aligned to the human reference genome, and single-nucleotide variants (SNVs) and small insertions and deletions (indels) were identified through variant calling. Quality control included assessment of sequencing coverage, genotype quality, missingness, sex concordance and relatedness. Genetic ancestry was inferred using principal component analysis, and ancestry-specific datasets were constructed for downstream GWAS

#### BioVU ancestry-specific GWAS

Ancestry-specific association analyses were performed using REGENIE v4^68^, adjusting for sex, age and the first five genetic principal components. In step 1, whole-genome ridge regression models were fitted using linkage-disequilibrium-pruned biallelic SNPs prepared with PLINK 2^69^. Linkage disequilibrium pruning used a 1,000-variant window, a 100-variant step and an r² threshold of 0.45. Variants were required to have a minor allele frequency (MAF) ≥ 1%, minor allele count ≥ 100, missingness ≤ 10% and Hardy–Weinberg equilibrium P ≥ 1 × 10⁻¹⁵. In step 2, association testing was performed for WGS-derived variants using leave-one-chromosome-out predictions from step 1 to account for polygenic effects and relatedness. Variants with association P < 0.01 were retested using approximate Firth logistic regression to reduce small-sample bias and improve the reliability of effect estimates for imbalanced case–control data. Following phenotype ascertainment and quality control, the final analyses included 2,581 CRC cases and 144,746 controls of European ancestry and 347 CRC cases and 26,048 controls of African ancestry.

#### GWAS datasets and phenotype harmonization

We combined the BioVU^27^ results with published and biobank CRC GWAS summary statistics: our previous multi-ancestry CRC meta-analysis^1^, the Million Veteran Program^28^, FinnGen release 13^29^, the UCLA ATLAS Community Health Initiative^30^, the Michigan Genomics Initiative Freeze 6^31^ and the Taiwan Precision Medicine Initiative^32^. In the additional electronic-health-record-linked biobanks, colon cancer cases were identified using ICD-9-CM and ICD-10-CM codes mapped to phecode 153.2. FinnGen cases were defined by the C3_COLON endpoint, based on Finnish Cancer Registry ICD-O-3 topography and morphology codes and harmonized ICD-8, ICD-9 and ICD-10 codes (**Supplementary Table 5**).

#### GWAS meta-analysis

GWAS summary statistics were harmonized to GRCh38 using UCSC LiftOver^70^, excluding unmapped variants and retaining the most significant record for duplicate variants. Analyses were restricted to autosomal variants with minor allele frequency (MAF) >1%. Inverse-variance-weighted fixed-effects meta-analyses were performed using METAL^71^, with genomic control applied to individual studies and meta-analysis results. Between-study heterogeneity was assessed using Cochran’s Q test and the I² statistic. The European-ancestry meta-analysis included 99,260 cases and 1,119,512 controls, whereas the multi-ancestry meta-analysis additionally incorporated African-, East Asian-, admixed American-and mixed-ancestry samples, totaling 132,261 cases and 1,752,398 controls (**Supplementary Table 5**).

### Cell-type-specific TWAS analysis

#### Building scTWAS GReX models

Model training was restricted to participants with matched germline genotype and single-cell expression data, with one biospecimen retained per participant. For the integrated VUMC normal-colon dataset, COLON MAP^24^ specimens were required to match the specimen metadata and be classified as normal mucosa; adenoma and serrated specimens were excluded. When multiple eligible specimens were available for a participant, one was selected according to the following anatomical-site priority: ascending colon, terminal ileum, cecum, descending colon, rectum, sigmoid colon, hepatic flexure, splenic flexure and transverse colon; ties were resolved by order of appearance in the expression matrix. Genotype data were matched to COLON MAP participants using unique participant identifiers, ensuring a one-to-one correspondence between participants and genotype samples.

For each cell type, gene-level expression counts were aggregated across cells from the same donor to generate pseudobulk expression profiles. Aggregated counts were converted to transcripts per million (TPM) and log₂-transformed. Expression values were then quantile-normalized across donors and subjected to a gene-wise inverse normal transformation. To account for potential confounding, normalized expression was adjusted for age, sex, the first five genotype principal components and probabilistic estimation of expression residuals (PEER) factors^72^. The number of PEER factors was determined by the number of donors in each training cohort following GTEx recommendations: 15 factors for the polyp and VUMC cohorts and 45 factors for the Wellcome Sanger IBDverse cohort. Residualized expression values were used as phenotypes for subsequent GReX model training.

Following the sTF-TWAS framework^23^, candidate genetic predictors were restricted to variants overlapping cell-type-specific accessible chromatin regions identified from single-cell ATAC-seq data. Variants were required to have a minor allele frequency (MAF > 0.05) and a Hardy–Weinberg equilibrium test P value >1 × 10⁻⁶. For each gene, eligible variants within ±1 Mb of the transcription start site were used to train an elastic-net regression model (α = 0.5), following the PrediXcan framework^6^. Predictive performance was evaluated as the coefficient of determination (*R*²) between predicted and observed expression across held-out samples using three repeated tenfold cross-validation. Models with cross-validated *R*² > 0.01 were retained. The same criterion applied to normal-colon and polyp models; for normal colon, a gene was carried forward for a given cell type if the model met this threshold in either training cohort.

#### scTWAS association testing

Cell-type-specific GReX models were tested against the European-ancestry and multi-ancestry CRC meta-analyses using the summary-statistics-based association framework of S-PrediXcan^73^ as implemented for sTF-TWAS^23^. The *Z_g_* score, representing the risk between gene expression changes of *g* and colorectal cancer risk, was calculated following the equation (1)

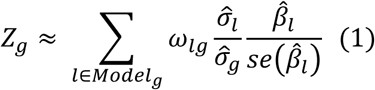

In Eq (1), *w_lg_* represents the predictive weight of variant *l* on expression residuals of gene *g*. β^*_l_* and *se*(β^*_l_*) represents the association regression coefficient and its standard error from GWAS SS for variant *l*, respectively. σ^*_l_* and σ^*_g_* are the estimated variances of variant *l* and the predicted expression of gene *g*. For the two normal-colon training cohorts, gene-level P values were combined using the Cauchy combination test using ACAT package^74^. Statistical significance was defined within each cell type by Bonferroni correction according to the number of retained gene models. Prioritized genes were identified primarily from the European-ancestry analysis, with additional genes from the multi-ancestry analysis when they reached cell-type-specific Bonferroni-adjusted *P* < 0.05.

### Colocalization and SMR-HEIDI of scTWAS genes

For each significant gene-cell-type association, we assessed whether the cis-expression and CRC association signals were consistent with a shared causal variant. A cis region of ±500 kb around the transcription start site (GENCODE v26) was defined for each gene, and CRC GWAS summary statistics were harmonized with cell-type-specific cis-eQTL summary statistics from the cohort contributing the association. Alleles were aligned, strand-ambiguous variants were excluded and at least 50 overlapping variants were required.

Bayesian colocalization was performed with coloc^33^ using the approximate Bayes factor approach. CRC was treated as a case-control trait (European-ancestry meta-analysis: 99,260 cases and 1,119,512 controls; multi-ancestry meta-analysis: 132,261 cases and 1,752,398 controls), and expression was treated as a quantitative trait using the corresponding cohort sample size. Minor-allele frequencies were obtained from the cis-eQTL data, and default prior probabilities were used (*p*1 = 1 × 10⁻⁴, *p*2 = 1 × 10⁻⁴ and *p*12 = 1 × 10⁻⁵). A locus was considered colocalized at PP.H4 > 0.8.

SMR and the HEIDI heterogeneity test^34^ were performed over the same cis regions using SMR v1.4.2. For each gene, the variant with the smallest cis-eQTL *P* value among harmonized variants was used as the instrument and specified with --target-snp. HEIDI included 3-20 variants in linkage disequilibrium (*r*² = 0.05 − 0.9) with the instrument and with cis-eQTL *P* < 1.57 × 10⁻³. Variants with minor-allele frequency < 0.01 or an allele-frequency difference > 0.2 between datasets were excluded. Linkage disequilibrium was estimated from the genotypes of the cohort contributing the cis-eQTL signal, with a locus-specific PLINK reference restricted to harmonized variants. SMR support was defined as a Bonferroni-significant SMR association (adjusted *P* < 0.05) with HEIDI *P* ≥ 0.05. Associations satisfying either PP.H4 > 0.8 or the SMR-HEIDI criterion were considered to have evidence consistent with a shared causal signal.

### TF-risk gene regulatory network analysis

#### TF-risk gene co-expression analysis

We investigated potential regulatory relationships between CRC susceptibility genes and 51 TFs previously implicated in CRC susceptibility by our previous TF-focused association study^23^. Analyses used scRNA-seq data from normal colorectal mucosa from 278 participants in the Wellcome Sanger Institute IBDverse cohort and colorectal polyps from 84 participants in the VUMC HTAN COLON MAP cohort. Within each epithelial population, single-cell expression profiles were aggregated to donor-level pseudobulk profiles summarize the count values from cells, and Pearson correlations were calculated between each TF and susceptibility gene. In total, 27,846 TF–gene pairs were tested, comprising 20,247 pairs in normal mucosa and 7,599 in polyps. *P* values were adjusted for multiple testing using the Benjamini–Hochberg procedure. Significant positive TF–risk-gene co-expression was defined as Pearson *r* > 0.5 and FDR < 0.05. A total of 1,397 TF–gene pairs met these criteria, including 982 in normal mucosa and 415 in polyps, and were retained for downstream regulatory network analyses (**Supplementary Table 14**).

#### TF ChIP-seq analysis linking TFs to susceptibility genes

To identify TFs potentially regulating CRC susceptibility genes, we integrated GReX prediction models with TF ChIP-seq data. For each significant gene–cell-type association, variants included in the corresponding GReX model were ranked by their association P values in the CRC GWAS meta-analysis. European-ancestry GWAS results were used for associations significant in the European-ancestry analysis, whereas multi-ancestry GWAS results were used otherwise. The variant with the smallest GWAS P value was designated the GReX lead variant and used to infer potential TF–risk gene regulatory relationships. A TF–risk gene link was established when the lead variant overlapped a ChIP-seq peak for the corresponding TF.

#### Integration of TF-risk gene regulatory evidence and TF prioritization

To prioritize key TFs potentially regulating CRC susceptibility genes, we integrated two complementary measures within each tissue: (i) the number of unique susceptibility genes linked to each TF through co-expression and (ii) the number of susceptibility-gene lead variants overlapping the corresponding TF ChIP-seq peaks. Each measure was converted to a percentile rank across the 51 candidate TFs, with missing values assigned zero before ranking. The two percentile ranks were averaged to calculate a composite regulatory evidence score, reflecting both co-expression connectivity and TF occupancy at risk-associated variants. TFs were ranked separately in normal colorectal mucosa and adenomas, and the top 20 TFs from each tissue were selected for downstream regulatory network analyses.

#### Cell-type specificity of TF–susceptibility gene relationships

To identify epithelial populations enriched for TF-linked CRC susceptibility genes, we included cell types with at least ten tested susceptibility genes: seven in normal colorectal mucosa (STM, TAC, ABS, GOB, EE, TUF and CT; 397 gene–cell-type pairs) and six in polyps (STM, TAC, ABS, GOB, ASC and SSC; 140 gene–cell-type pairs). EE, TUF and CT were excluded from polyp analyses owing to insufficient gene coverage. For each of the 51 candidate TFs, enrichment of co-expressed susceptibility genes within each cell type was assessed relative to the pooled remaining cell types from the same tissue using a one-sided Fisher’s exact test, treating each gene–cell-type pair as an independent observation. In total, 357 TF–cell-type combinations were tested in normal mucosa and 306 in polyps. P values were adjusted separately for each tissue using the Benjamini–Hochberg procedure, with significant enrichment defined as FDR < 0.05 and at least three linked susceptibility genes (**Supplementary Table 17**). We selected two normal epithelial populations showing the strongest TF enrichment (ABS and GOB) and two polyp-associated populations (ASC and SSC) for network visualization. Within each population, up to six TFs with the smallest nominal enrichment P values (P < 0.05) and at least three linked susceptibility genes were selected. Susceptibility genes co-expressed with at least two selected TFs were retained; when fewer than five genes met this criterion, all linked susceptibility genes were included. For TFs lacking connections to the retained genes, the two most strongly co-expressed susceptibility genes were additionally incorporated. Network edges represented significant positive TF–gene co-expression (Pearson r > 0.5, FDR < 0.05), with TF ChIP-seq occupancy overlapping the corresponding GReX lead variant highlighted to indicate complementary regulatory evidence.

### Differential Expression Analyses of Omics and Spatial Data Across Colorectal Tumorigenesis: From Normal Mucosa to Adenoma and CRC

Unless stated otherwise, a gene-resource pair was considered directionally supported when the gene was differentially expressed or differentially abundant at FDR < 0.05, showed |log2FC| > 0.5, and changed in the direction predicted by the sign of the scTWAS Z score. Pairs meeting the significance and effect-size thresholds but changing in the opposite direction were classified as discordant; pairs not meeting these thresholds were classified as non-significant; and genes not measured by a resource were classified as not tested, so that absence of measurement was never treated as negative evidence. For genes prioritized in multiple cell-type models, the association with the smallest P value was selected to determine the representative direction of effect. Positive scTWAS Z scores indicated that higher genetically predicted expression was associated with increased CRC risk, whereas negative Z scores indicated an association with reduced CRC risk.

#### Pseudobulk differential expression analysis across tumorigenesis

Transcriptional support was assessed in the COLON MAP single-cell resource^16^, which spans normal colorectal epithelium, conventional adenoma, serrated polyp and carcinoma and therefore represents both the normal-adenoma-carcinoma and the normal-serrated polyp-carcinoma routes. Depending on cell type, analyses included up to 86 normal colon, 75 conventional adenoma and 46 serrated polyp specimens together with seven carcinomas (two MSI-H and five MSS). Raw counts were aggregated per epithelial cell type within each specimen to give donor-level pseudobulk profiles; specimens with at least 10 cells of a given cell type were retained and genes detected in at least 10% of retained pseudobulk samples were analysed. Data were handled with scanpy v1.11.5^75^ (Python v3.11.16) and differential expression was performed with PyDESeq2 v0.5.4^76^ using negative-binomial generalized linear models and Wald tests, with cohort of origin included as a covariate for contrasts spanning multiple cohorts. Log2 fold changes were shrunk with apeglm^77^, whereas test statistics and Benjamini-Hochberg-adjusted P values were taken from the unshrunken model. For the lesion-restricted models we evaluated support across 23 epithelial-stage transitions grouped into three stages. Complete-progression transitions compared each of the seven normal epithelial cell types with MSI-H and with MSS carcinoma cells, giving 14 contrasts. Early-stage transitions compared each of the seven normal-colon epithelial cell types with ASC or with SSC. Late-stage transitions compared ASC or SSC with MSI-H and MSS carcinoma cells. All contrasts were oriented as later versus earlier disease stage, so that positive log2FC indicates increased expression with progression. Support was defined as above.

#### TCGA analyses

TCGA-COAD bulk RNA-sequencing data were obtained from the Genomic Data Commons with TCGAbiolinks v2.38.0^78^ in R v4.5.0, using SummarizedExperiment v1.40.0 and DESeq2 v1.50.2^79^. STAR counts from primary tumour and solid-tissue-normal specimens were analysed, retaining one sample per patient per condition. Ensembl version suffixes were removed and duplicated identifiers collapsed by summing counts. Genes with at least 10 reads in at least five samples were retained, and differential expression between primary tumour and normal colon was estimated with DESeq2 using a negative-binomial generalized linear model with tumour-normal stages in the design, oriented as tumour versus normal.

#### CPTAC analyses

Protein-abundance data from the CPTAC colon adenocarcinoma cohort (University of Michigan dataset) were retrieved using the Python package cptac (v1.5.14). Protein features were mapped to gene symbols, with multiple features mapping to the same gene averaged. Differential protein abundance between tumour and adjacent-normal tissues was assessed using a paired two-sided *t*-test when at least five matched pairs were available, or Welch’s two-sided *t*-test when fewer than five matched pairs but at least three samples per group were available. Analyses were performed on log-transformed abundances, with log2 fold changes (log2FC) calculated as tumour minus normal. *P* values were adjusted using the Benjamini–Hochberg procedure, with a false discovery rate (FDR) threshold of 0.05.

#### GeoMx analyses

Spatial transcriptional support was evaluated using published NanoString GeoMx data^42^ from 19 stage T1 colorectal cancers, comprising independent Cancer Transcriptome Atlas and Whole Transcriptome Atlas cohorts. Spatial regions representing normal mucosa, adenoma, tumour core and invasive front were segmented into PanCK-positive epithelial and PanCK-negative stromal compartments. We used the study authors’ differential-expression results from gene-wise linear mixed-effects models of log2-normalized expression as a function of tissue region, with patient-specific random intercepts and tissue-region slopes. Contrasts were oriented as later versus earlier disease stage. Nominal P-values were adjusted by the Benjamini–Hochberg method across the prespecified scTWAS genes within each experiment, compartment and contrast. A gene was considered spatially supported when it showed FDR < 0.05 in the normal-versus-tumour-core epithelial contrast, the expression change was concordant with its scTWAS direction, and no significant epithelial result in either GeoMx cohort showed an opposing direction. Nonsignificant opposing estimates did not disqualify a gene. Normal-versus-adenoma and adenoma-versus-tumour-core contrasts, invasive-front contrasts and stromal results were analyzed descriptively but were not used to define the primary spatially supported set.

#### CosMx analyses

CosMx data were obtained from Buissant des Amorie et al.^80^ (Zenodo 10.5281/zenodo.17671259, Nanostring_CosMx_exp2.zip), comprising 1,257,627 quality-controlled cells from 11 early colorectal cancers profiled across two slides. The 21 published cell-type annotations were grouped into carcinoma, normal epithelial, oncofetal, fibroblast or stromal, immune and other populations for visualization. Gene expression counts were normalized at the single-cell level. Analyses were restricted to prioritized susceptibility genes represented on the 6,175-gene CosMx panel. Patients containing both carcinoma and normal or undefined epithelial populations were included in paired analyses. For each gene, normalized expression was averaged within each patient and epithelial compartment, excluding groups containing fewer than 50 cells. Expression differences between carcinoma and normal or undefined epithelial compartments were assessed using a paired Wilcoxon signed-rank test across patients with eligible paired measurements. Effect sizes were calculated as the mean within-patient expression difference (carcinoma minus normal or undefined epithelium). Directional concordance was defined as agreement between the sign of this difference and the corresponding scTWAS Z score. P values were adjusted across the prioritized genes represented on the panel using the Benjamini–Hochberg procedure. For spatial visualization, representative genes were selected based on nominal significance in paired analyses (P < 0.05), CRC relevance or druggability, and availability of per-cell expression data. Cells expressing at least one selected gene were retained together with a random subset of the remaining cells to preserve tissue context. All quantitative summaries, including cell counts and proportions, were calculated using the complete dataset rather than the visualization subset.

### Drug-target annotation and functional enrichment analyses

#### Drug-target identification

Druggable CRC susceptibility genes and associated compounds were identified by integrating drug-protein interaction data from DrugBank^37^, ChEMBL^38^, TTD^39^ and Open Targets^40^. For each compound, we extracted commercial names, highest clinical trial phase and approved indications, and classified susceptibility genes as druggable if they encoded targets of approved or clinical-stage compounds. Mechanisms of action were categorized as decreasing target activity (inhibitors, antagonists or negative allosteric modulators) or increasing target activity.

#### Annotation of Established CRC-Associated Genes

Established cancer genes were identified by matching the 347 prioritized CRC susceptibility genes against approved gene symbols in the COSMIC Cancer Gene Census v103 (GRCh38; 763 genes, including 590 Tier 1 and 173 Tier 2)^81^. Census gene synonyms were additionally examined, yielding no further matches. Enrichment of Cancer Gene Census membership among the 63 druggable susceptibility genes relative to the remaining 284 susceptibility genes was assessed using a two-sided Fisher’s exact test.

#### KEGG pathway enrichment

Pathway over-representation was evaluated using human KEGG annotations retrieved from the KEGG REST API^82^. The reference universe comprised 9,290 genes annotated to at least one KEGG pathway, including 53 of the 63 druggable genes and 147 of the 347 prioritized susceptibility genes. We evaluated 119 pathways containing at least two druggable genes using two reference backgrounds: all 9,290 KEGG-annotated human genes, assessed by one-tailed hypergeometric tests, and the 147 KEGG-annotated susceptibility genes, assessed by one-sided Fisher’s exact tests. Both analyses tested over-representation, with Benjamini-Hochberg correction applied separately across the 119 pathways within each comparison.

### iLINCS connectivity analysis

To identify compounds capable of reversing genetically predicted CRC susceptibility-associated transcriptional changes, we performed connectivity analysis using the iLINCS^44^ LIB5 large-intestine perturbation library, comprising 16,767 compound perturbation signatures. We evaluated 43 prespecified compounds targeting prioritized CRC susceptibility genes, matching compounds between the drug annotation table and iLINCS^44^ using PubChem compound identifiers (CIDs) rather than compound names. Each compound perturbation signature was compared with 18 cohort-level cell-type TWAS signatures containing more than 200 genes overlapping with the iLINCS^44^ reference using normalized connectivity scores (NCS)^83^. Negative NCS values indicated opposing transcriptional responses, such that genes with genetically predicted risk-increasing expression were downregulated by the compound, whereas genes with risk-decreasing expression were upregulated. For each perturbation signature, NCS values were averaged across the 18 cell-type TWAS signatures. A compound-level mean NCS was then calculated across all available library signatures for that compound.

To account for variation in the number of perturbation signatures available per compound, we constructed a size-matched empirical null distribution by randomly sampling 5,000 sets of library signatures, with each set containing the same number of signatures as the compound under evaluation. The compound-level mean NCS was compared against the corresponding empirical null distribution using a one-sided test for negative connectivity. Empirical *P* values were adjusted across the 43 compounds using the Benjamini-Hochberg procedure, with an FDR threshold of 0.05. Compounds were ranked by their observed mean NCS, and the null distribution for each compound was summarized by its mean ± 2 standard deviations (s.d.). Compounds represented by only one perturbation signature were retained for completeness but interpreted cautiously because their mean connectivity estimates could not be assessed for consistency across independent library signatures.

### DepMap dependency analysis

Dependency data. Gene-level CRISPR knockout fitness data were obtained from the Cancer Dependency Map (DepMap) through the Open Targets Platform GraphQL API (release 26.09, depMapEssentiality). Records were restricted to the DepMap intestine tissue category, comprising 63 colorectal adenocarcinoma cell lines, and deduplicated by gene symbol and DepMap identifier. Dependency was quantified using Chronos gene-effect scores, where 0 indicates no detectable proliferation effect and −0.5 approximates the moderate effect of knocking out a common essential gene. For each gene, the median score across evaluable cell lines was used as the dependency measure.

#### Gene sets

Four gene sets were evaluated against the same cell-line panel: (1) 347 prioritized scTWAS risk genes, of which 274 had scores available in more than 90% of cell lines; (2) 202 CRC consensus susceptibility genes supported by at least two of ten published studies^1–3,8–14^, of which 175 were evaluable; (3) 52 Tier 1 CRC-associated driver genes from the COSMIC Cancer Gene Census v103 (GRCh38)^81^, all evaluable; and (4) 466 randomly selected background genes. CRC driver genes were identified by systematically classifying tumour-type annotations in the census somatic and germline fields, excluding non-colorectal intestinal and gastrointestinal annotations. The background was derived from 500 genes randomly sampled from a 2,000-gene regulatory and protein-protein interaction network background, excluding prioritized risk genes and Hart core-essential (CEGv2) and non-essential (NEGv1) reference genes^84^. Genes overlapping any focal set were subsequently removed.

#### Dependency classification and statistical analysis

A gene was classified as a broadly shared dependency if its median Chronos gene-effect score was below −0.5 and at least 90% of evaluable cell lines had scores below −0.5. Gene-set differences were assessed using the Kruskal-Wallis test, followed by two-sided Mann-Whitney U tests for six pairwise comparisons, with Holm correction for multiple testing. Individual genes were treated as the units of analysis. Effect sizes were summarized as differences in group medians and Hodges-Lehmann location shifts. Because focal sets partially overlapped, pairwise comparisons were repeated after excluding shared genes from the smaller set. Sensitivity analyses additionally evaluated four alternative CRC driver-gene definitions based on census tier, somatic annotations and fusion-only stages. Analyses were performed using Python 3.11, NumPy 2.4, pandas 2.3, SciPy 1.16 and statsmodels 0.14.

### Arrayed CRISPR-Cas9 screen

#### CRC cell lines and culture

Human colorectal cancer cell lines HCT116, RKO, and SW480 were obtained from the American Type Culture Collection (ATCC, Manassas, VA, USA) and maintained in RPMI-1640 medium containing 2 mM L-glutamine, 1 mM sodium pyruvate, 10 mM HEPES, and 1.5 g/L sodium bicarbonate, supplemented with 10% fetal bovine serum (FBS) and penicillin-streptomycin (100 U/mL penicillin and 100 µg/mL streptomycin). Cells were cultured at 37°C in a humidified atmosphere containing 5% CO₂.

#### Candidate Gene Selection and CRISPR–Cas9-Mediated Knockout

Candidate genes were prioritized for functional validation based on two criteria: (i) significant scTWAS associations indicating that genetically predicted gene expression was associated with increased CRC risk and (ii) prior evidence from published studies supporting their functional relevance to CRC development or tumorigenesis. This strategy prioritized genetically implicated genes with complementary biological evidence for experimental evaluation of their effects on CRC cell viability. To produce stable cell lines, SW480, RKO and HCT116 cells were transduced with Dharmacon Edit-R Inducible Lentiviral Cas9 particles at low MOI (1-10), selected with blasticidin, and expanded as a pool. Cas9 expression was induced with doxycycline treatment 24 hr prior to transfection. Edit-R predesigned synthetic crRNA were annealed with tracrRNA for each gene and prepared in 384-well cell plates at Xpmol per well. DharmaFECT reagent was added to wells for 20 min followed by addition of Cas9-expressing RKO or HCT116 cells with a final concentration of crRNA:tracrRNA of 50 nM. Two independently designed Edit-R non-targeting crRNA (#1 and #2) and lethal crRNA (#1) were also included on each plate as negative and positive controls, respectively. Cells were incubated for 72 hours to allow for gene-editing and effects on cell viability.

#### Live/dead cell assay

At 72 hours post-transfection, Hoechst 33342 and propidium iodide (PI) were added to each well at 1:5,000 dilution and allowed to incubate at 37 degrees for 20 minutes. Stained cell plates were then imaged on the Molecular Devices ImageXpress Micro XLS using a 4x objective to capture the entire well and both DAPI and TxRed channels. Total nuclei and PI+ cells were segmented and quantified using MetaXpress software and the Multiwavelength Cell Scoring application module. Each gene was compared to the mean of non-targeting controls. Experiments were repeated in triplicate.

#### Statistical analysis of the arrayed CRISPR–Cas9 screen

Cell viability was calculated as the percentage of PI-negative cells among total nuclei at 72 hours after transfection. For each target gene, viability was normalized to the corresponding non-targeting crRNA controls within each experiment. Differences in normalized viability between target-gene knockout and control conditions were evaluated using a two-sided paired Student’s t-test across three independent biological replicates (n = 3), with experimental replicate as the matching factor. P values were adjusted across target genes within each cell line using the Benjamini–Hochberg procedure, with FDR < 0.05 considered statistically significant. Results were reported as mean ± standard deviation (s.d.).

## Statistics and reproducibility

All donors and specimens meeting the prespecified cohort-specific quality-control criteria were included. Donors or patients were treated as independent biological replicates in single-cell, pseudobulk and spatial analyses. Exclusion criteria, including genotype and sample quality, minimum cell counts, gene detection thresholds and plate-level quality control, are described in the corresponding Methods sections. No donors were excluded after analysis. Statistical tests were two-sided unless otherwise specified. One-sided tests were used for prespecified directional hypotheses, including TF enrichment across cell types (**Supplementary Table 17**), KEGG pathway over-representation among druggable genes (**Supplementary Table 23**) and empirical tests for negative drug connectivity. Cancer Gene Census enrichment was assessed using a two-sided Fisher’s exact test. The HEIDI test was used to assess heterogeneity, with P ≥ 0.05 indicating no statistically significant evidence of heterogeneity. Multiple-testing correction was performed using the Bonferroni method for scTWAS associations within each cell type, the Benjamini–Hochberg procedure for differential expression, co-expression, enrichment, drug connectivity and spatial analyses, and the Holm method for pairwise gene-dependency comparisons. Statistical significance was defined as adjusted P < 0.05 unless otherwise specified. Arrayed CRISPR–Cas9 experiments were performed in three independent biological replicates. For each target gene, normalized cell viability was compared with matched non-targeting controls using a two-sided paired Student’s t-test, followed by Benjamini–Hochberg correction across target genes within each cell line. All replicates passing plate-level quality control were included. Randomization and blinding were not implemented for the computational analyses; experimental randomization and blinding were not reported. Exact P values are provided where computationally representable.

## Code availability

Code for model training, association testing, network construction and figure generation is available at https://github.com/XingyiGuo/scTWAS, together with environment specifications and the analysis workflow. GitHub repository in Zenodo and obtain a DOI.

## Data Availability

The European-ancestry and multi-ancestry CRC GWAS meta-analysis summary statistics generated in this study will be deposited in the GWAS Catalog, https://github.com/XingyiGuo/scTWAS_in_CRC/tree/main/data/GWAS_SS.Previously published CRC GWAS summary statistics for European populations are available in the GWAS Catalog under accession GCST90129505. IBDverse genotype data are available from the European Genome-phenome Archive under accession EGAD00001016069 under the terms of the originating study^21^.

Single-cell RNA-sequencing data from the Colorectal Molecular Atlas Project (COLON MAP) are available through the NCI Human Tumor Atlas Network Data Portal https://humantumoratlas.org. Gut Cell Atlas single-cell data are available under GSE266546. Single-cell ATAC-sequencing and Individual-level genotype data from the VUMC HTAN COLON MAP and VUMC Gut Cell Atlas cohorts are available [GSE123456]; the Stanford HTAN colorectal atlas scATAC-seq data are available under GSE201336. The GeoMx and CosMx spatial datasets analysed here were obtained from Zenodo (10.5281/zenodo.17671259)^80^.

Reference and annotation resources used were: TCGA-COAD transcriptomes from the Genomic Data Commons with the R package named TCGAbiolinks; CPTAC-COAD proteomes via the cptac Python package; Cancer Dependency Map (DepMap) through the Open Targets Platform GraphQL API (release 26.09, depMapEssentiality); GENCODE v26 (GRCh38) (https://www.gencodegenes.org/human/release_26.html); ChEMBL (https://www.ebi.ac.uk/chembl/); TTD (https://db.idrblab.net/ttd/); DrugBank (https://go.drugbank.com/); Open Targets (https://www.opentargets.org/); COSMIC Cancer Gene Census v103 (https://cancer.sanger.ac.uk/census); MSigDB v7.3; and iLINCS (https://www.ilincs.org/). Source data for all main figures are provided with the paper. A consolidated inventory of all datasets and reference resources analysed in this study, with accessions, versions and access conditions, is provided in **Supplementary Table 1**.

## Acknowledgements

This research was primarily supported by US National Institutes of Health grants R37CA227130, R01CA269589 and R01CA297582 to X.G, R01DK103831 and U54CA274367 to K.S.L. Additional support was provided by The Leona M. and Harry B. Helmsley Charitable Trust through grant G-1903-03793 to Vanderbilt University Medical Center, which supported single-cell RNA sequencing and genotyping, and by the US Department of Veterans Affairs Merit Review grants I01CX002662 and I01CX002473 (the latter to K.T.W.).

We thank the Million Veteran Program staff, researchers and, above all, the veterans whose participation made this research possible. We acknowledge the participants and investigators of FinnGen. We gratefully acknowledge the support of the Institute for Precision Health, the participating patients of the UCLA ATLAS Precision Health Biobank, the UCLA David Geffen School of Medicine, the UCLA Clinical and Translational Science Institute (UL1TR001881) and UCLA Health. We acknowledge the Michigan Genomics Initiative participants, AI and Digital Health Innovation at the University of Michigan, the University of Michigan Medical School Central Biorepository and Advanced Genomics Core for specimen storage, management, processing and distribution, and the Center for Statistical Genetics in the Department of Biostatistics at the School of Public Health for genotype curation, imputation and management. We thank the participants and investigators of the Taiwan Precision Medicine Initiative, and Academia Sinica and the cooperating medical centers across Taiwan for their support in establishing and maintaining the cohort datasets.

## Author contributions

X.G. and K.S.L. conceived and designed the study. Q.L., C.L. and X.G. developed the study methodology. Q.L., C.L. and L.L. developed the computational tools and performed the statistical and bioinformatics analyses. Q.L., C.L., L.L., J.W. and J.A.B. conducted the investigations, performed validation analyses and curated the data. F.Y., Qi.S., J.C., S.X., Y.W., Qu.S., C.Y., E.J., Y.C., Q.C., W.W., X.-O.S., G.G., B.S., M.J.S., Z.Y., L.A.C., K.K.C., C.E., J.A.B., U.P., K.T.W., W.Z. and K.S.L. contributed data, samples and other research resources. Q.L. and C.L. prepared the figures and visualizations. Q.L. and X.G. drafted the initial manuscript. All authors reviewed and edited the manuscript and approved the final version. X.G. supervised the study. X.G., K.S.L. and K.T.W. acquired funding to support the research.

---

**Extended Data Fig. 1.**
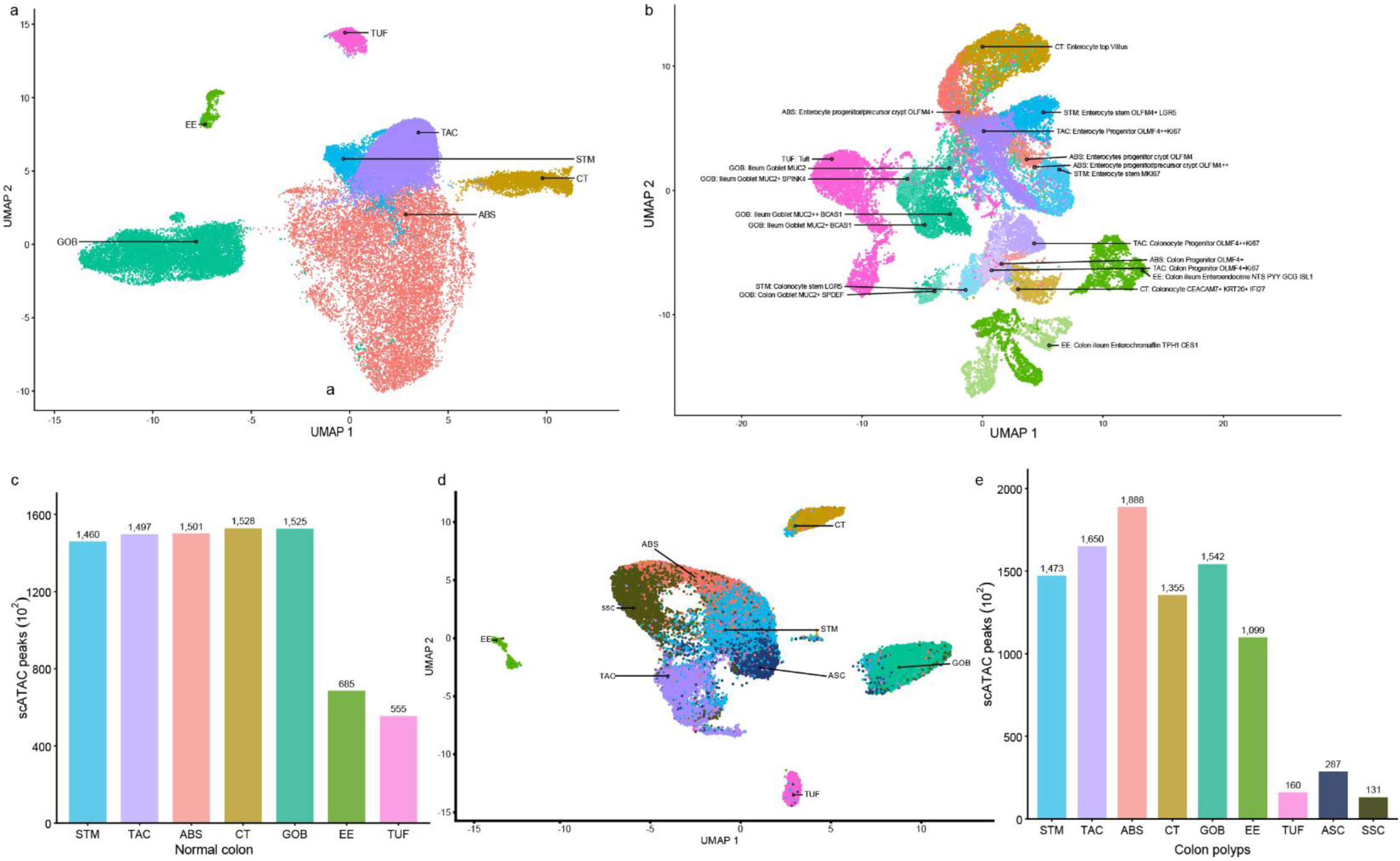
Cell-type composition, chromatin accessibility and model performance in normal colon. **a**, UMAP of the VUMC HTAN and Gut Cell Atlas normal-colon scRNA-seq data, coloured by the seven epithelial populations used for model training: absorptive (ABS), crypt-top (CT), enteroendocrine (EE), goblet (GOB), stem (STM), transit-amplifying (TAC) and tuft (TUF). **b**, UMAP of the Wellcome Sanger IBDverse normal-colon data, labelled with the published fine-grained cell annotations and the population each was harmonized to for this study (Supplementary Table 3). **c**, Number of reproducible scATAC-seq peaks called in each population after merging the VUMC and Stanford HTAN peak sets, in units of 10² (Supplementary Table 5). **d**, UMAP of the VUMC HTAN COLON MAP polyp scRNA-seq data, coloured by the nine epithelial populations used for model training: lesion-restricted adenoma-specific (ASC), serrated-polyp-specific (SSC) states and the seven shared with normal colon (ABS, CT, EE, GOB, STM, TAC and TUF). **e**, Number of reproducible scATAC-seq peaks called in each population after merging the VUMC and Stanford HTAN peak sets, in units of 10²; the TUF, ASC and SSC annotations derive from the VUMC dataset alone (Supplementary Table 5).

**Extended Data Fig. 2.**
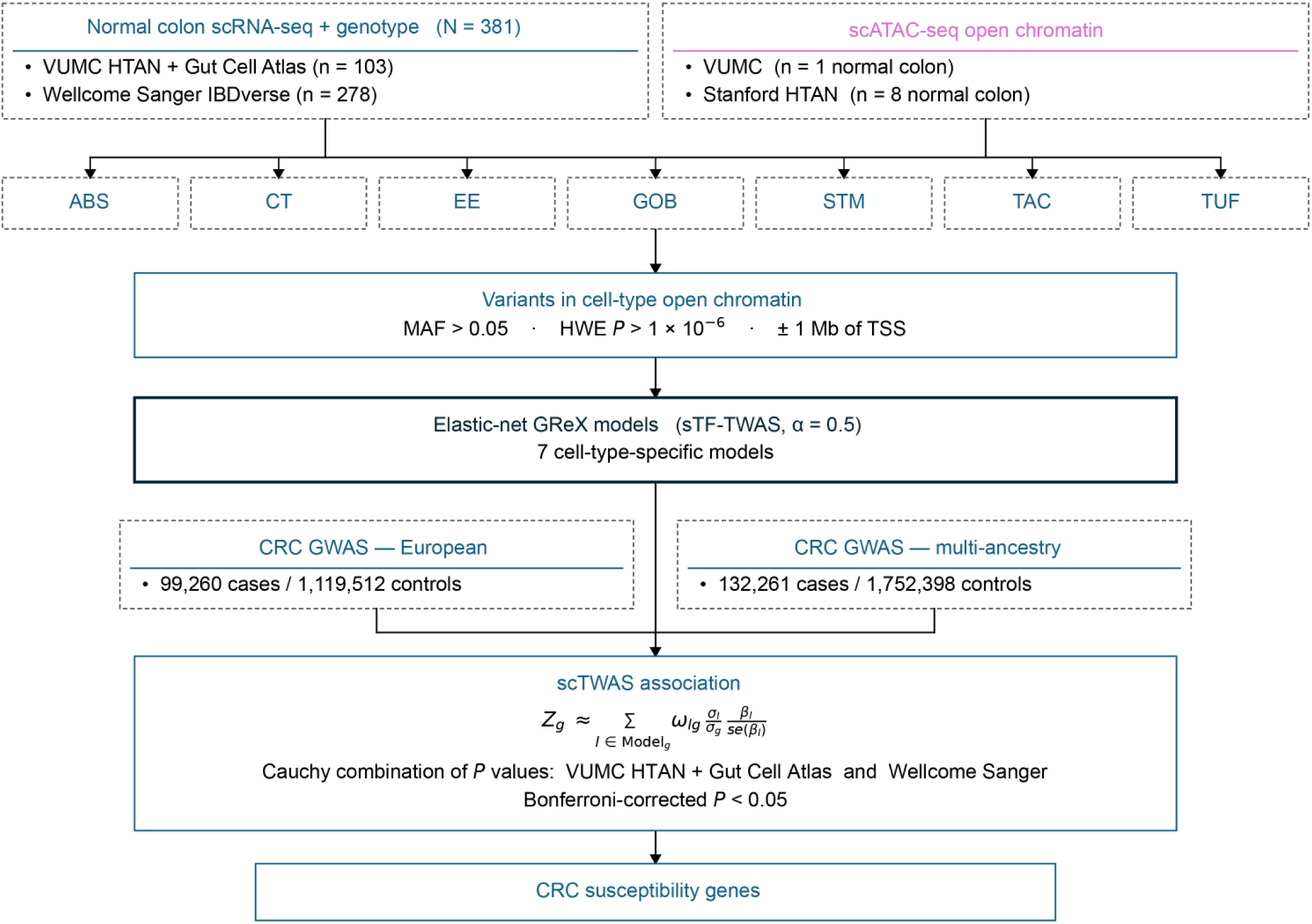
Overview of the scTWAS workflow for normal colorectal epithelium cells. Cell-type-specific genetically regulated expression (GReX) models were trained using matched germline genotype and scRNA-seq data from the VUMC and Wellcome Sanger normal colon donors. Candidate regulatory variants were prioritized using cell-type-specific chromatin accessibility profiles, followed by elastic-net model training. The resulting models were integrated with European-ancestry and multi-ancestry CRC GWAS summary statistics, with association P values across training cohorts combined using the Cauchy combination test. Colocalization and SMR–HEIDI analyses provided additional genetic support for prioritized CRC susceptibility genes.

**Extended Data Fig. 3.**
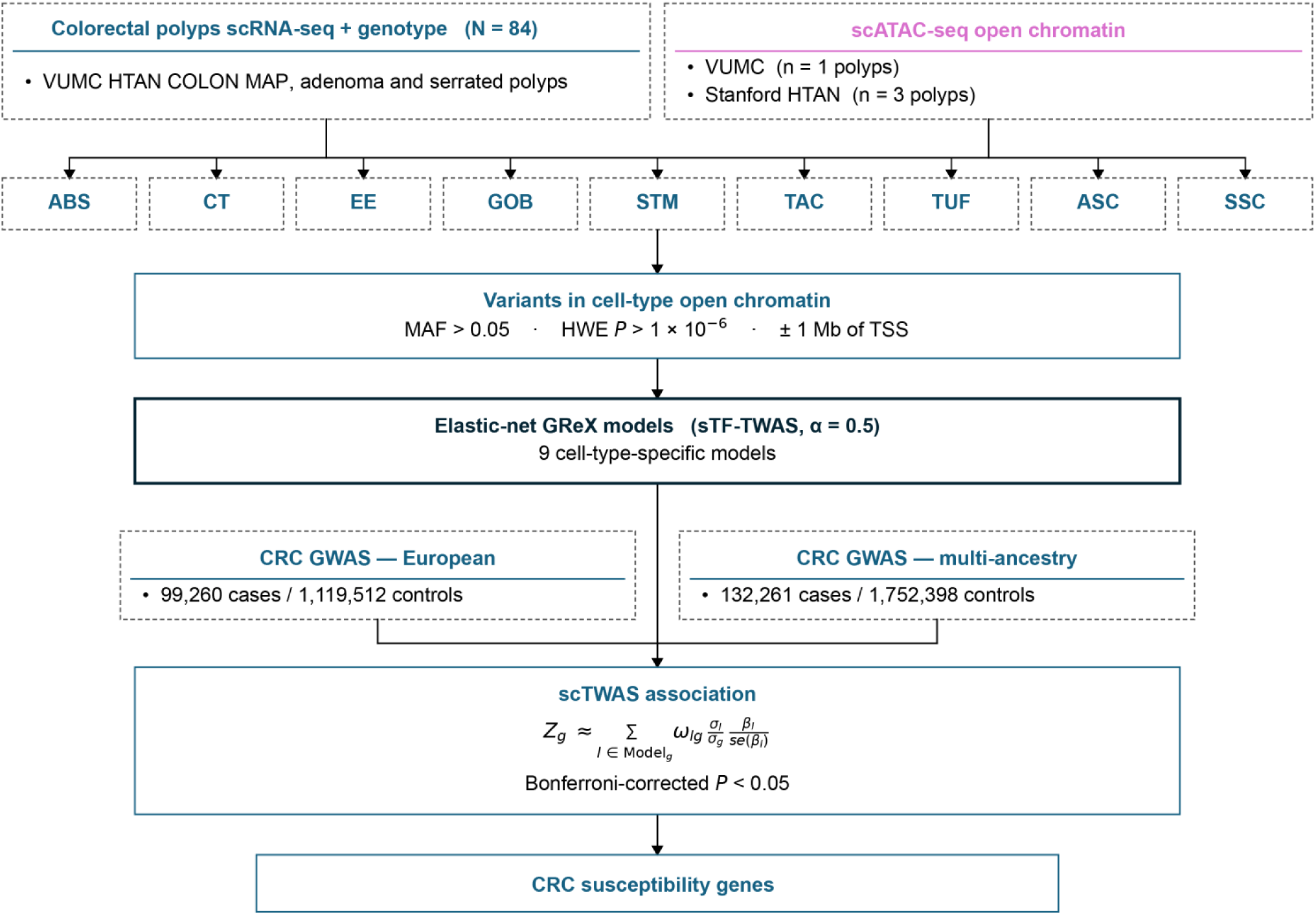
Overview of the scTWAS workflow for normal colorectal polyps cells. Cell-type-specific genetically regulated expression (GReX) models were trained using matched germline genotype and scRNA-seq data from the VUMC polyps donors. Candidate regulatory variants were prioritized using cell-type-specific chromatin accessibility profiles, followed by elastic-net model training. The resulting models were integrated with European-ancestry and multi-ancestry CRC GWAS summary statistics, with association P values across training cohorts combined using the Cauchy combination test. Colocalization and SMR–HEIDI analyses provided additional genetic support for prioritized CRC susceptibility genes.

**Extended Data Fig. 4.**
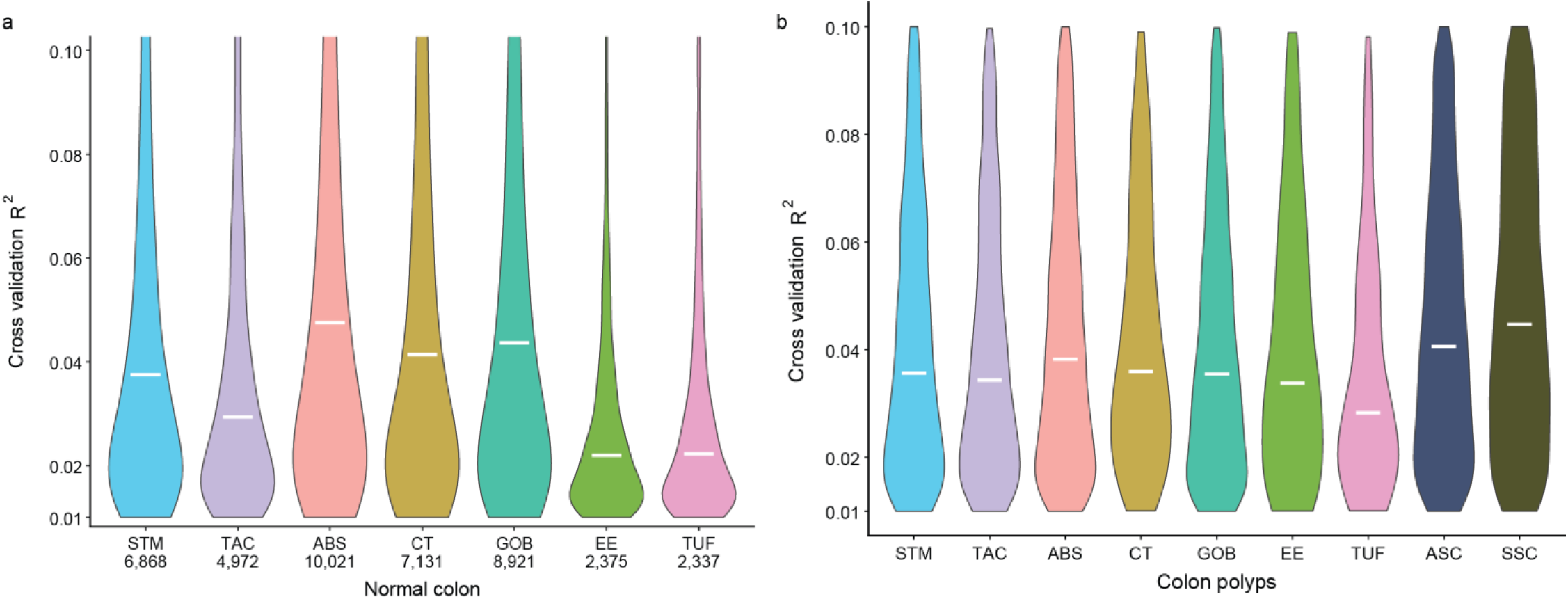
Predictive performance of cell-type-specific GReX models in normal colorectal epithelium and colorectal polyps. **a**, Distribution of cross-validated R² values for GReX models across seven normal-colon epithelial populations from the VUMC HTAN and Wellcome Sanger cohorts. Numbers below the cell-type labels indicate the number of gene models. **b**, Distribution of cross-validated R² values for GReX models across nine colorectal polyp epithelial populations from the VUMC cohort. White horizontal bars indicate median R² values. Distributions are displayed within the R² range of 0.01-0.10.

**Extended Data Fig. 5.**
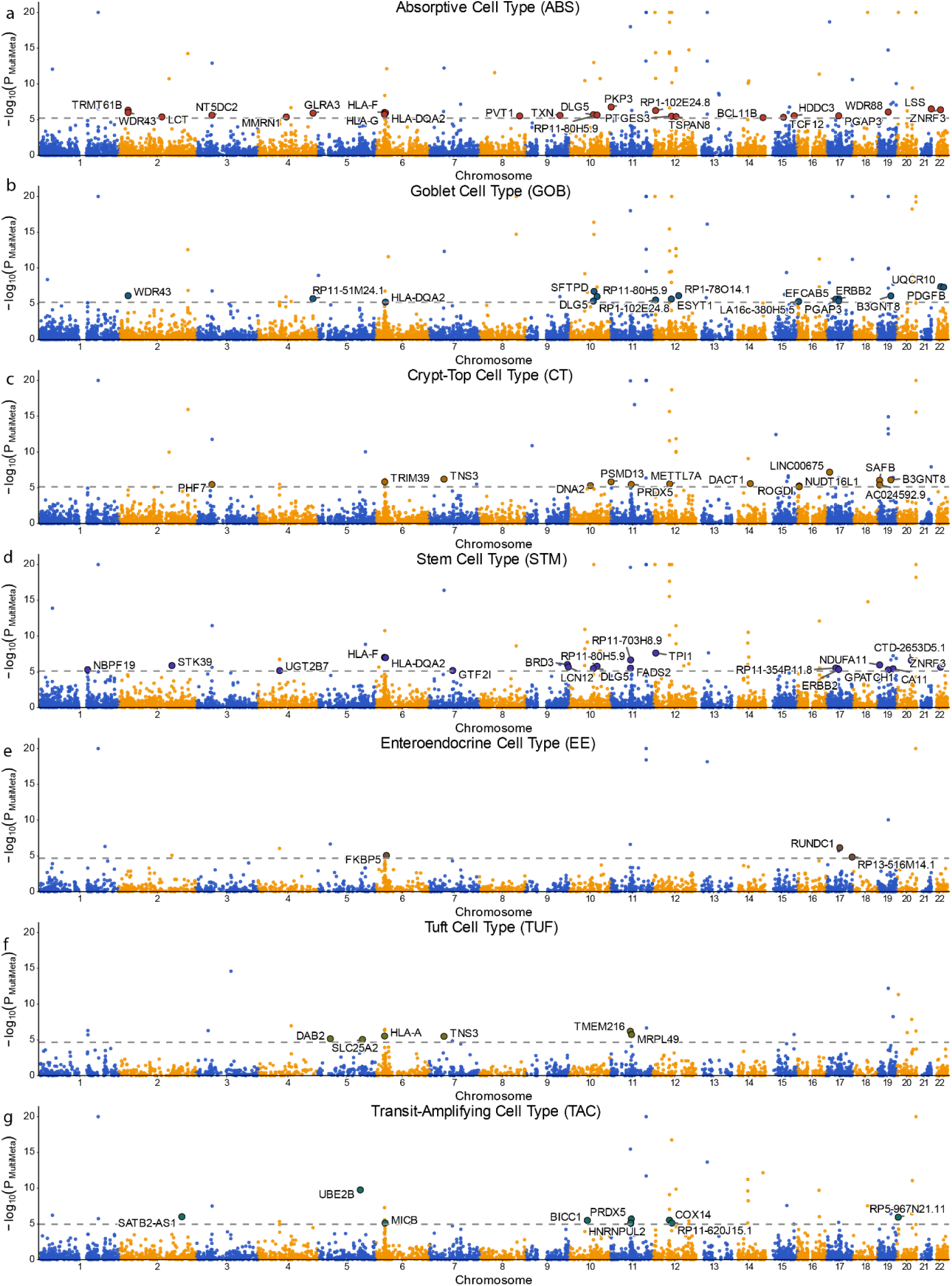
Multi-ancestry scTWAS associations in normal colon. a-g, Genome-wide distribution of associations for absorptive (a), goblet (b), crypt-top (c), stem (d), enteroendocrine (e), tuft (f) and transit-amplifying (g) cells. Each point is one gene, plotted by chromosomal position against the negative log10 of its Cauchy-combined multi-ancestry P value, with alternating colours distinguishing consecutive chromosomes. The dashed line marks the cell-type-specific Bonferroni threshold. Enlarged, labelled points are the significant associations contributed by the multi-ancestry analysis beyond those already significant in the European-ancestry analysis shown in Figure 2.

**Extended Data Fig. 6.**
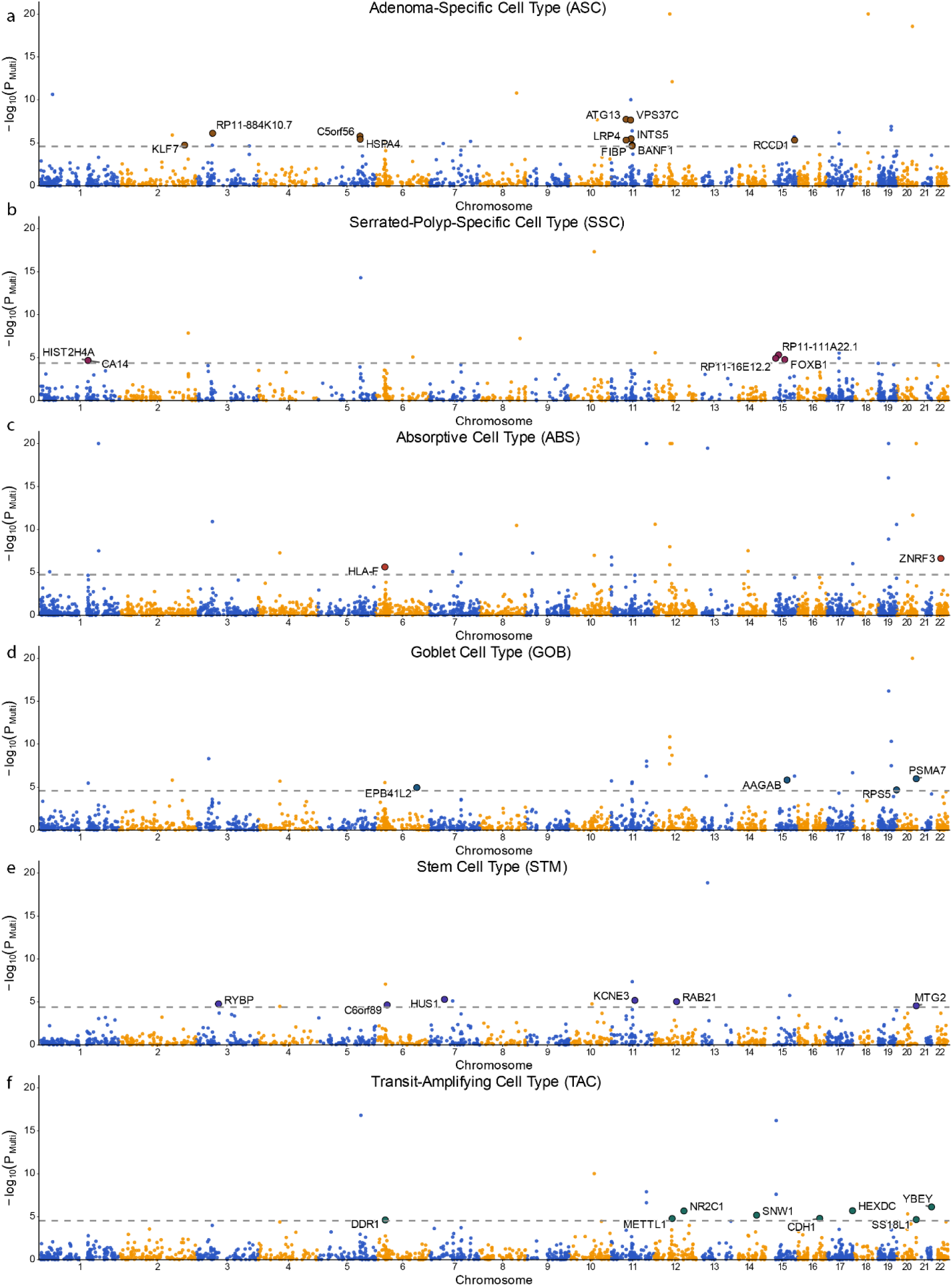
Multi-ancestry scTWAS associations in colorectal polyps. **a-f**, Genome-wide distribution of associations for adenoma-specific (a), serrated-polyp-specific (b), absorptive (c), goblet (d), stem (e) and transit-amplifying (f) cells. Axes, chromosome shading and the dashed Bonferroni threshold are as in Extended Data Fig. 3. Enlarged, labelled points are the significant associations contributed by the multi-ancestry analysis beyond those already significant in the European-ancestry analysis shown in Figure 3.

**Extended Data Fig. 7.**
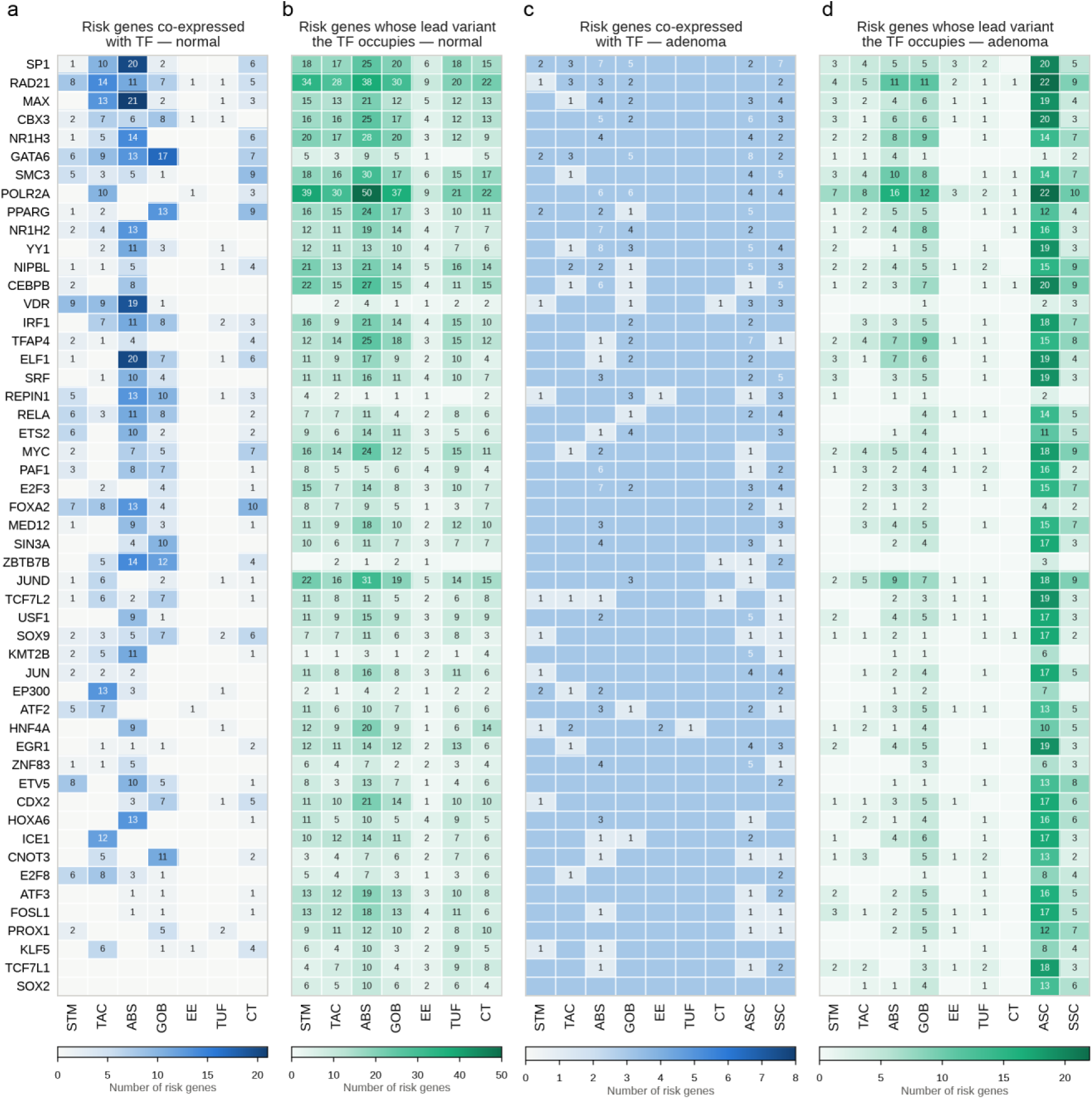
Cell-type distribution of two regulatory evidence layers linking CRC susceptibility-associated transcription factors to risk genes. a–d, For each of the 51 CRC susceptibility-associated transcription factors (rows) and epithelial populations (columns), heat maps show the number of prioritized CRC risk genes supported by TF–gene co-expression (a,c) or TF occupancy at the GWAS lead variant linked to the risk gene (b,d). Co-expression was defined as Pearson *r* > 0.5 with FDR-adjusted *P* < 0.05 across donor-level pseudobulk profiles. TF occupancy was defined by overlap of colon TF ChIP–seq peaks with the corresponding GWAS lead variant; thus, cell-type differences in b and d reflect the cell-type-specific sets of prioritized risk genes rather than cell-type-specific ChIP–seq profiles. a,b, Normal colorectal epithelium across seven populations (STM, TAC, ABS, GOB, EE, TUF and CT). c,d, Colorectal polyp (ADSER) epithelium across nine populations, additionally including the lesion-restricted ASC and SSC states. Numbers indicate risk-gene counts; blank cells indicate no supported risk genes. Colour scales are panel-specific and therefore comparable within, but not across, panels.

**Extended Data Fig. 8.**
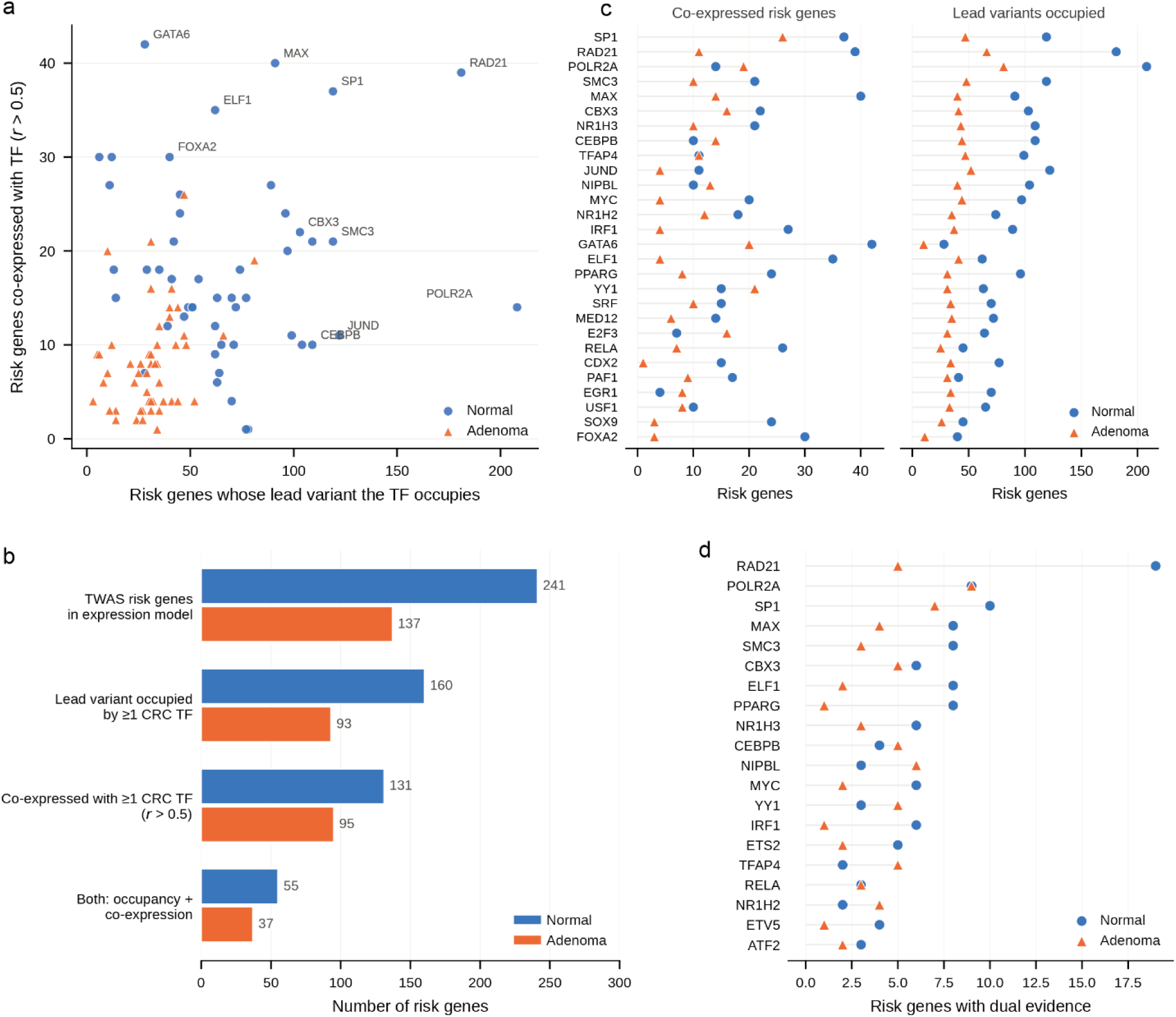
Occupancy and co-expression provide complementary regulatory evidence. **a**, Relationship between TF occupancy and TF–risk gene co-expression. Each point represents one TF in one tissue and plots the number of risk genes whose GWAS lead variant is occupied by the TF (x axis) against the number of risk genes co-expressed with that TF at Pearson *r* > 0.5 (y axis), shown for normal colon (circles) and adenoma and serrated lesions (triangles). Labeled TFs represent extreme values on either axis. Within cell types, the two layers were only weakly correlated (Spearman ρ = −0.21 to 0.37). b, Number of risk genes supported by each evidence layer in each tissue: genes represented in the cell-type-specific expression models (241 in normal colon, 137 in polyps), genes whose GWAS lead variant is occupied by at least one CRC susceptibility TF (160 and 93), genes co-expressed with at least one such TF at Pearson *r* > 0.5 (131 and 95), and genes supported by both layers (55 and 37). c, Occupancy and co-expression evidence for the 28 key TFs, defined as the union of the 20 highest-ranked TFs in normal colon and the 20 highest-ranked TFs in polyps, of which 12 rank in the top 20 of both tissues (Methods). TFs are ordered by the sum of their ranks for the two evidence layers in the two tissues. For each tissue, the left panel gives the number of co-expressed risk genes and the right panel the number of risk-gene lead variants occupied by the TF. d, Number of risk genes supported by both occupancy and co-expression evidence for each TF, shown for the 20 TFs with the largest numbers of dual-evidence genes in each tissue. These 20 TFs are selected on dual-evidence count and are not the same set as the 28 key TFs in c; ETS2, ATF2 and ETV5 appear in d only.

**Extended Data Fig. 9.**
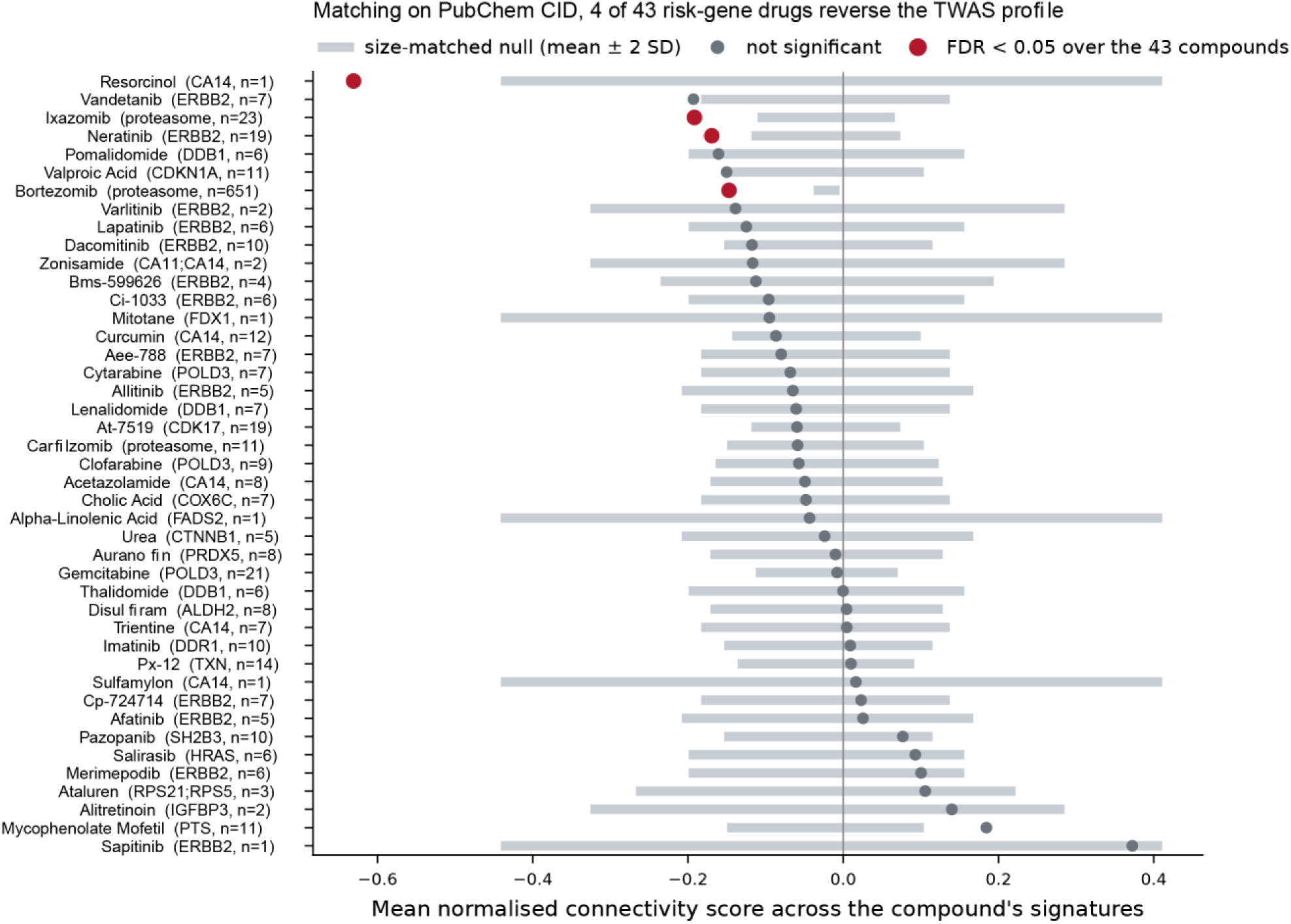
Compound-induced reversal of cell-type-specific CRC susceptibility signatures. Connectivity analysis of 43 prespecified compounds targeting prioritized CRC susceptibility genes against 18 cohort-level cell-type TWAS signatures using the iLINCS LIB5 large-intestine perturbation library (16,767 signatures). Compounds are ranked by their mean normalized connectivity score (NCS), calculated by first averaging across the 18 cell-type signatures and then across all available perturbation signatures for each compound. Points indicate observed mean NCS; horizontal bars represent the mean ± 2 standard deviations (s.d.) of size-matched empirical null distributions generated from 5,000 random signature sets. Red points indicate significant negative connectivity (Benjamini–Hochberg FDR < 0.05 across 43 compounds, one-sided empirical test); grey points indicate nonsignificant results. Negative NCS indicates reversal of the genetically predicted CRC risk-associated expression profile. Compound labels identify the targeted risk gene or target class and the number (n) of contributing perturbation signatures. Proteasome inhibitors target the proteasome complex, including PSMA7, PSMD3 and PSMD13. Compound identities were harmonized using PubChem compound identifiers (CIDs). Resorcinol (n = 1) was retained but interpreted cautiously. Per-compound statistics are provided in Supplementary Table 24.

